# Multi-Omics Prognostic Marker Discovery and Survival Modeling: A Case Study on Pan-Cancer Survival Analyses in Women’s Cancers

**DOI:** 10.1101/2025.01.08.25320212

**Authors:** Ricky Nguyen, Fatemeh Vafaee

## Abstract

Survival analysis is essential for predicting patient outcomes and guiding personalized cancer treatments. While multi-omics data offers valuable insights, its high dimensionality complicates analysis and clinical application. Many studies still rely on the traditional Cox proportional hazards model, with limited exploration of alternative survival algorithms or robust feature selection methods. Few frameworks effectively integrate features across multiple omics modalities. To address these issues, we developed PRISM (PRognostic marker Identification and Survival Modelling through Multi-omics Integration), a comprehensive framework designed to improve survival predictions and identify key prognostic markers. PRISM systematically compares various feature selection methods and survival models and employs a robust pipeline that selects features from single-omics data, integrating them through feature-level fusion and multi-stage refinement. Applied to TCGA data for Breast Invasive Carcinoma (BRCA), Cervical Squamous Cell Carcinoma and Endocervical Adenocarcinoma (CESC), Ovarian Serous Cystadenocarcinoma (OV) and Uterine Corpus Endometrial Carcinoma (UCEC), PRISM demonstrated that integrating DNA methylation, miRNA, and copy number variation data provided complementary information, significantly outperforming other modality combinations in three cancers (BRCA: C-index 0.77; CESC: 0.80; UCEC: 0.76). Pan-cancer analysis further revealed shared oncogenic pathways and therapeutic targets. PRISM provides a scalable, generalizable solution for multi-omics integration, advancing cancer research and precision medicine.

**Key Points:**

- **Advancing Multi-Omics Survival Modelling** – PRISM introduces a robust computational framework that enhances survival predictions and identifies key prognostic markers by systematically integrating multi-omics data.
- **Rigorous Feature Selection and Benchmarking** – PRISM systematically evaluates multiple feature selection techniques and survival models using a voting-based approach to improve methodological robustness.
- **Optimized and Cost-Effective Signature Panel** – By employing recursive feature elimination (RFE), PRISM minimizes the number of selected biomarkers, improving cost-effectiveness while maintaining high predictive accuracy for clinical applications.
- **Comprehensive Multi-Omics Integration** – PRISM enables the fusion of features across diverse omics modalities using single-stage and two-tier refinement strategies, optimizing the final prognostic signature for maximum predictive power.
- **Scalability and Clinical Relevance** – PRISM is generalizable beyond TCGA datasets, making it applicable to large-scale multi-omics studies with broad clinical and biomedical implications for personalized medicine and cancer research.

## Introduction

Cancer’s complex pathophysiology is shaped by diverse genetic, environmental, and molecular factors, leading to considerable variability in patient outcomes even within the same cancer types, which complicates treatment strategies [1,2]. High-throughput molecular profiling technologies, such as next-generation sequencing and mass spectrometry, have become fundamental in precision medicine, enabling comprehensive analysis of DNA, RNA, and proteins to discover biomarkers. However, relying on single omics data provides only partial insights into the intricate mechanisms of cancer, potentially missing critical biomarkers and therapeutic opportunities [3,4]. The heterogeneity of cancer, reflected in its diverse subtypes and molecular profiles, requires an integrated approach. Combining multiple omics data types is crucial for gaining a holistic understanding of cancer biology and enabling personalized treatment strategies [5]. The vast array of omics data—spanning genomics, transcriptomics, proteomics, metabolomics, and epigenomics—offers a comprehensive view of cancer biology, with immense potential to identify novel biomarkers and improve clinical outcomes [6,7]. However, multi-omics data poses challenges like high dimensionality, data imbalance, noise, missing values, and heterogeneity, complicating robust analysis and biomarker discovery [5]. To address these challenges, integrative machine learning methods such as multimodal learning, ensemble strategies, and network-based approaches are increasingly crucial in advancing biomarker discovery and clinical outcome models for disease diagnosis, prognosis, and treatment response monitoring [8–12]. In this context, survival analysis is pivotal, helping researchers and clinicians evaluate factors influencing patient outcomes over time. Predicting survival beyond critical milestones, such as five-year survival, informs treatment decisions and enhances our understanding of cancer progression [3]. This growing recognition of the importance of multi-omics survival modeling has spurred numerous studies in the past decade.

Yuan et al. [13] conducted a pioneering study by integrating multiple omics data—including somatic copy-number alterations (SCNA), DNA methylation, mRNA, miRNA, and protein expression—to predict patient survival across four cancer types from The Cancer Genome Atlas (TCGA) project. This work established the value of multi-omics integration for survival analysis. Building on this, Zhu et al. [14] systematically evaluated the prognostic value of various omics types —clinical data, mRNA, SCNA, DNA methylation, miRNA—across 14 cancers using a kernel machine learning approach. They found gene and miR1NA expression to have the strongest predictive power. Liu et al. [15] focused on multi-omics variable selection combined with Cox-regression model for cancer prognosis prediction. Subsequently, Chai et al. [2] integrated multi-omics data to derive representative features, which were then utilized in a Cox proportional hazards (Cox-PH) model for enhanced survival prediction. Later, Chaudhary et al. [16] employed an autoencoder to integrate DNA methylation, miRNA expression, and RNA-Seq data, selecting survival-associated features using univariate Cox-PH analysis and identifying robust subgroups in hepatocellular carcinoma (HCC). Similarly, Cheerla et al. [17] utilized a deep learning approach with multi-modal representations to enhance cancer prognosis and Cox-PH for outcome prediction. Hao et al. [18] extended deep learning methodologies by proposing a gene- and pathway-based deep neural network (DNN) where the Cox-PH model served as the output layer, embedding multi-omics features for survival prediction. Tong et al. [19] further advanced the field by introducing multi-view learning with autoencoders, integrating multi-omics data through complementary and consensus principles. Their approach demonstrated how DNA methylation and miRNA expression offer both unique and shared prognostic insights, with the integrated model significantly outperforming single-omics methods.

While studies emphasize the value of multi-omics integration for cancer survival modeling, few systematically evaluate survival algorithms beyond Cox Proportional Hazards (CoxPH) or assess feature selection alongside diverse models. A unified framework consolidating top-performing features across omics layers is also lacking. To address these gaps, we developed PRISM (PRognostic marker Identification and Survival Modelling through Multi-omics Integration), an end-to-end framework for survival prediction and prognostic biomarker identification. PRISM systematically analyzes TCGA multi-omics data, including gene expression, DNA methylation, miRNA expression, and copy number variations. It employs a comprehensive feature selection and survival modeling pipeline, using statistical and machine learning techniques to extract key biomarkers, integrate them via two fusion methods, and evaluate their predictive power. PRISM benchmarks feature selection methods, such as univariate/multivariate Cox filtering and Random Forest importance, alongside survival models including CoxPH, ElasticNet, GLMBoost, and Random Survival Forest [3]. Through cross-validation, bootstrapping, ensemble voting, and recursive feature elimination (RFE), it enhances robustness and minimizes signature panel size without compromising performance. Designed for adaptability beyond TCGA, PRISM offers a complete pipeline from data retrieval to functional analysis and visualization. It improves survival prediction, aiding patient stratification, personalized treatment, and precision medicine. To demonstrate its utility, PRISM was applied to women-related cancers from TCGA—BRCA, OV, CESC, and UCEC—major contributors to female cancer burden [20,21]. Despite distinct molecular profiles, these cancers share pathways influencing progression and therapy response [22,23]. Pan-cancer analyses further reveal shared oncogenic drivers and therapeutic targets. While applied here to women-related cancers, PRISM is a general-purpose framework extendable to any cancer type, advancing precision oncology and improving patient outcomes [24].

## Methods

### Omics Modalities and Cancers

Multi-omics data were obtained from The Cancer Genome Atlas (TCGA) (https://portal.gdc.cancer.gov/) using the TCGAbiolink R package. Table 1 summarizes the number of samples available for each omics data type across the studied cancers, focusing on four women-related cancers: Breast Invasive Carcinoma (BRCA), Ovarian Serous Cystadenocarcinoma (OV), Cervical Squamous Cell Carcinoma and Endocervical Adenocarcinoma (CESC), and Uterine Corpus Endometrial Carcinoma (UCEC). For each cancer type, data were collected on gene expression (GE), copy number variations (CNV), DNA methylation (DM), miRNA expression (ME), and clinical variables, including patient vital status and the number of days to death or last follow-up, which were integrated for survival analysis. To ensure consistency, only samples with complete data across all five categories were included. Age was considered as a potential confounding variable, and using the BRCA dataset, we divided the age range (26–89 years) into three quantile-based groups (Supplementary Figure 4). A Kruskal-Wallis test revealed no significant differences (p > 0.05) in the expression levels of biomarkers such as miR-22 and miR-150 across these groups. Kaplan-Meier survival analysis (Supplementary Figure 5) stratified by age also showed no significant survival differences, suggesting age does not confound the relationship between these biomarkers and survival. These findings, detailed in the Supplementary Material, support the exclusion of age as a primary variable in this study. While similar analyses can be conducted for other factors, our focus remains on omics-based integration, with demographic and exposure-related variables considered as potential additional data modalities.

**Table 1.** Overview of the TCGA sample counts for each omics data type across various cancer types. The "Clinical" column represents samples with available survival data, while "Common" indicates patient samples with complete data across all five omics types.

| TCGA Cancer Types | Gene Expression (GE) | Copy Number Variation (CNV) | DNA Methylation (DM) | MiRNA expression (ME) | Clinical | Common |
| --- | --- | --- | --- | --- | --- | --- |
| **BRCA:**<br>Breast Invasive Carcinoma | 1111 | 1050 | 1096 | 1096 | 1098 | 739 |
| **OV:**<br>Ovarian Serous Cystadenocarcinoma | 421 | 557 | 582 | 490 | 608 | 397 |
| **CESC:**<br>Cervical Squamous Cell Carcinoma and Endocervical Adenocarcinoma | 304 | 294 | 307 | 307 | 307 | 291 |
| **UCEC:**<br>Uterine Corpus Endometrial Carcinoma | 553 | 536 | 438 | 545 | 560 | 421 |

Feature extraction for each omics modality was performed using TCGA bioinformatics pipelines. Pre-processed multi-omics data were retrieved from the GDC Portal via the *TCGAbiolinks* R package. GE data were normalized to FPKM using the TCGA mRNA pipeline, encompassing over 60,000 features, including isoforms and non-coding RNAs. CNV data were derived from "Gene Level Copy Number Scores," indicating gene-level gains and losses. DM data consist of beta values (0–1) from Illumina 450K/27K assays, where 0 signifies no methylation and 1 indicates full methylation. ME data were quantified using a modified TCGA pipeline developed by BCGSC.

### Data pre-processing

Omics data were structured in tabular form, with samples as rows and features as columns. Only samples labeled "01" (Primary Solid Tumor) were retained. GE features were aligned with gene names and matched to CNV data, averaging multiple signals per gene within a sample. Clinical survival data were integrated into each matrix. Features and samples with >20% missing values in GE, CNV, and ME data were removed, followed by filtering for highly variable features. A log transformation, log(X+1), was applied (X = FPKM for GE, RPM for ME). DM data were reduced to 27k CpG sites for pan-cancer comparisons, ensuring compatibility with cancers like OV that only had 27k data. Samples/features with >50% missing values were removed, and remaining gaps were imputed using the mean. For ME, we retained miRNAs present in >50% of samples (value >0) and >10% (value >1), per Zhao et al. (2020) [1].

Pre-processed TCGA data were then passed through our feature selection pipeline to identify the final feature set for each omics, which was evaluated using predictive survival models. Before developing the pipeline, we assessed filter methods against a no-selection baseline (Figure 1). *Supplementary Table 5* details the machine learning algorithms, filter methods, R packages, and hyperparameters used.

**Figure 1.**
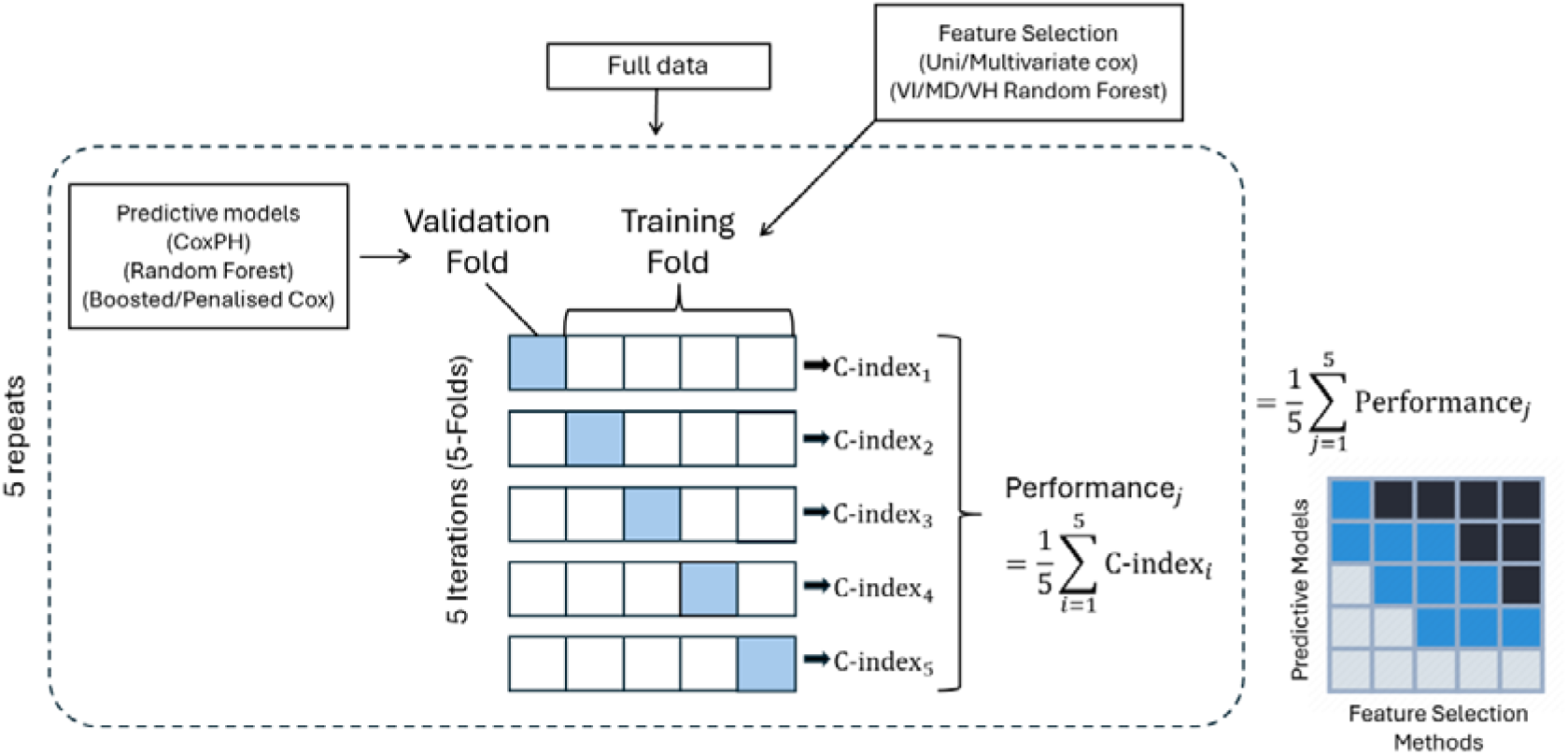
Evaluation of Individual Filter Methods Pipeline. Workflow for evaluating each filter method independently. Cross-validation is employed to assess the performance of each method, and the results are summarized in a heatmap displaying the mean performance of each filter method when paired with various predictive models

Figure 1 illustrates the evaluation process for each feature selection method. We applied 5 repeats of 5-fold cross-validation, splitting the data into five folds, with four for training and one for validation. Each filter method was applied to the training set, selecting features based on thresholded coefficients and importance scores. For the univariate and multivariate Cox methods, features were selected if they were significant (p < 0.05) and had non-zero coefficients. For the random survival forest method, features with an importance score > 0 were selected. The performance of these selected features was assessed using four survival machine learning algorithms, evaluating the concordance index (C-index) on the validation set. The C-index measures model discrimination, ranging from 0.5 (random guessing) to 1 (perfect prediction) [1]. This process was repeated 5 times, resulting in 25 evaluations per feature selection method and survival model. The final performance of each method was determined by averaging the C-index values.

### Feature Selection Pipelines

We developed two feature selection pipelines, one that utilizes cross-validation and another that uses bootstrapping. Figure 2(A) illustrates our cross-validation (CV) pipeline. We begin with a 70:30 training-test split, using the training set for feature selection. The training set is divided into 5 folds, with one-fold held out each time. Each filter method is applied to the remaining folds to identify important features. This process is repeated 5 times with 5-fold CV, resulting in 25 iterations. Features are ranked by their occurrence in the selected sets, with a maximum possible occurrence of 100 (25 iterations × 4 filter methods). Features selected at least 50% of the time are included in the final set. This final set is then evaluated on the test set to ensure no information leakage. The second feature selection method, the bootstrapping pipeline (Figure 2(B)), involves bootstrapping 70% of the data and applying each filter method. The common features selected across all methods form a bootstrapped feature set, repeated 100 times. We track the frequency of each feature’s occurrence, selecting those that appear at or above the 70th quantile. This approach identifies highly predictive features while accounting for data variability. To evaluate both pipelines, we test the final selected features using the separate test set. The training set, filtered to include only the selected features, is used to train predictive models, and performance is assessed on the independent test set. This ensures unbiased evaluation and robust assessment of predictive performance.

**Figure 2.**
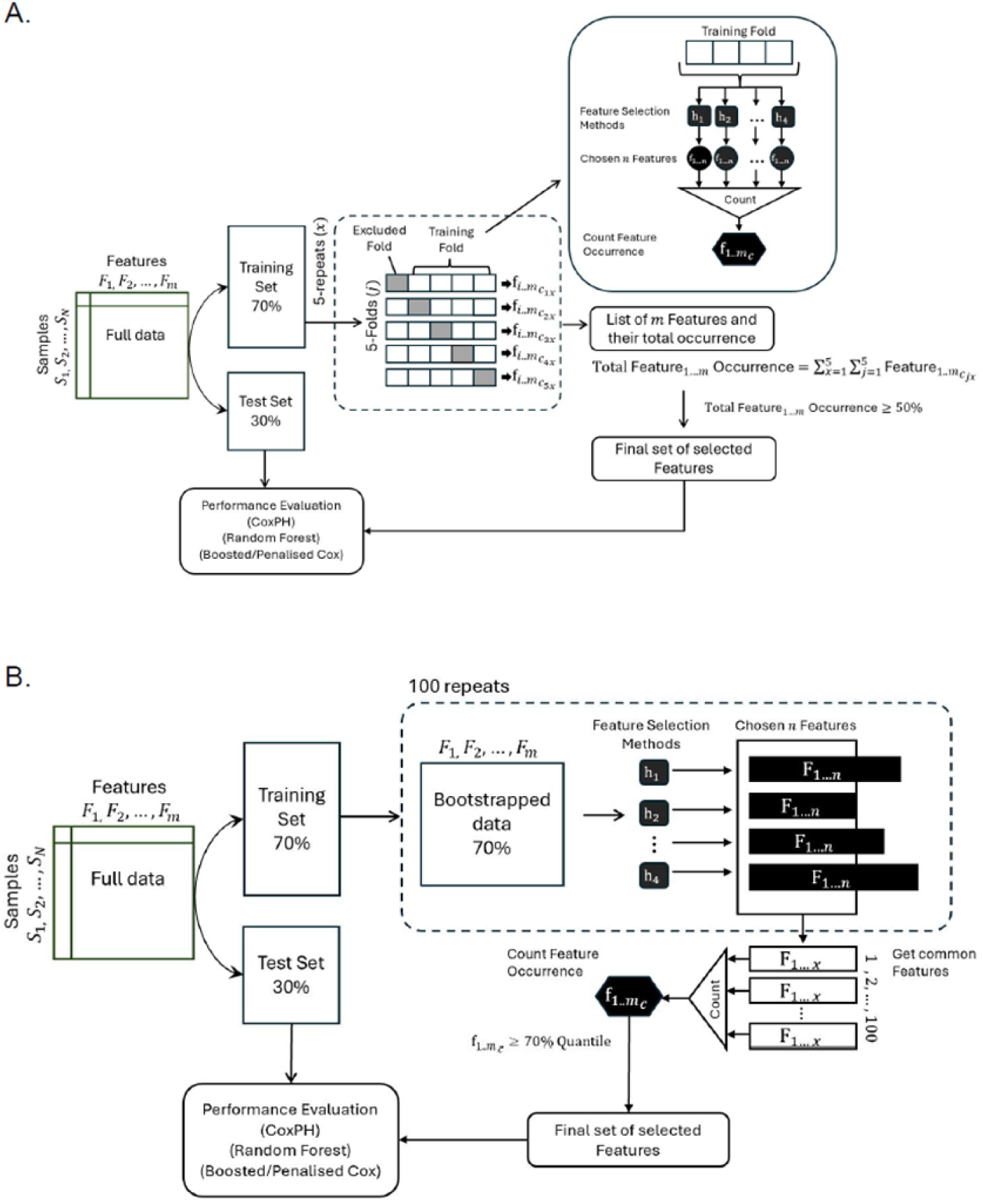
Feature Selection Pipelines. **A**.CV Feature selection pipeline. Apply a cross-validation process combined with a voting technique to identify optimal features. Selected features are determined based on occurrence across the chosen filter methods, enhancing the robustness of feature selection. **B.** Bootstrapping Feature selection pipeline. Employs bootstrapping to improve the reliability of selected features by resampling the data across numerous iterations and applying the same voting technique

### Multi-Omics pipeline

Following feature selection for each omics modality, we integrate multi-omics data using feature-level fusion, refining the signature panel to enhance predictive performance while ensuring clinical feasibility (Figure 3). To achieve this, we employ two refinement strategies. In **One-Stage Refinement**, selected features from each omics modality are first concatenated, and Recursive Feature Elimination (RFE) is applied to identify the most predictive features. In contrast, **Two-Stage Refinement** applies RFE at the single-omics level before fusion, then again at the multi-omics level, ensuring that only the most informative features are retained at each stage. This sequential approach enhances model robustness, minimizes overfitting, and improves overall predictive accuracy. To determine the optimal combination of omics modalities for survival prediction, we systematically evaluate all possible integrations, similar to Tong et al. (2020) [19], who used autoencoders for feature fusion. We assess model performance using various machine learning methods, including Cox Proportional Hazards, ElasticNet, Random Forest (RF), and GLMBoost. Ultimately, RF is selected for multi-omics integration due to its superior predictive accuracy (Figure 4, Table 2).

**Figure 3.**
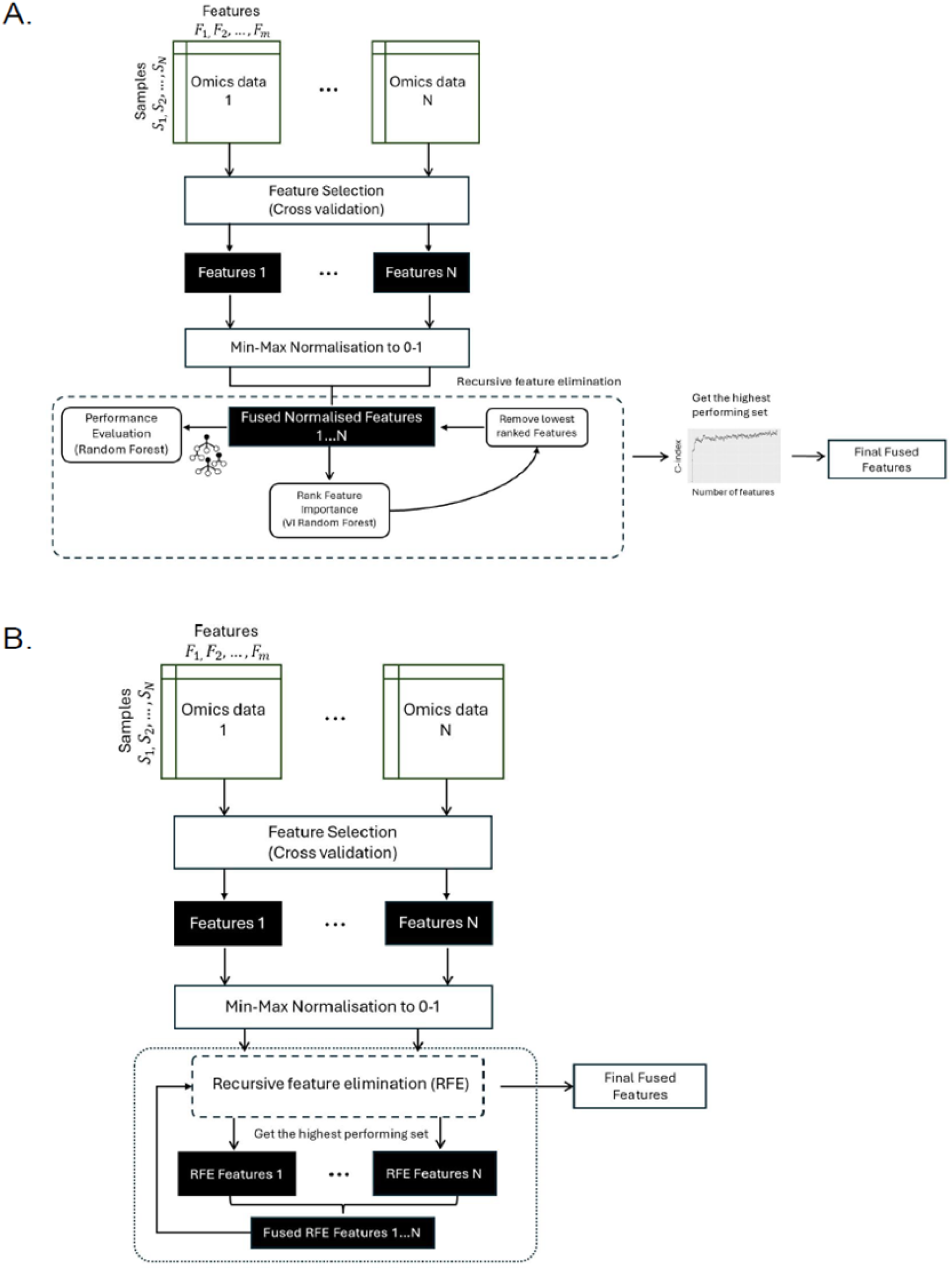
Multi-omics pipelines. **A.** One-Stage Refinement integration pipeline, where two modalities are combined at the feature level. Recursive Feature Elimination (RFE) is then applied to reduce dimensionality, further refining the feature set to enhance model performance. **B.** Two-Stage Refinement integration pipeline, where individual modalities first undergo Recursive Feature Elimination (RFE) to reduce dimensionality. The reduced feature sets are then fused together, followed by an additional round of RFE on the fused data to obtain the final set of multi-omics features

**Figure 4.**
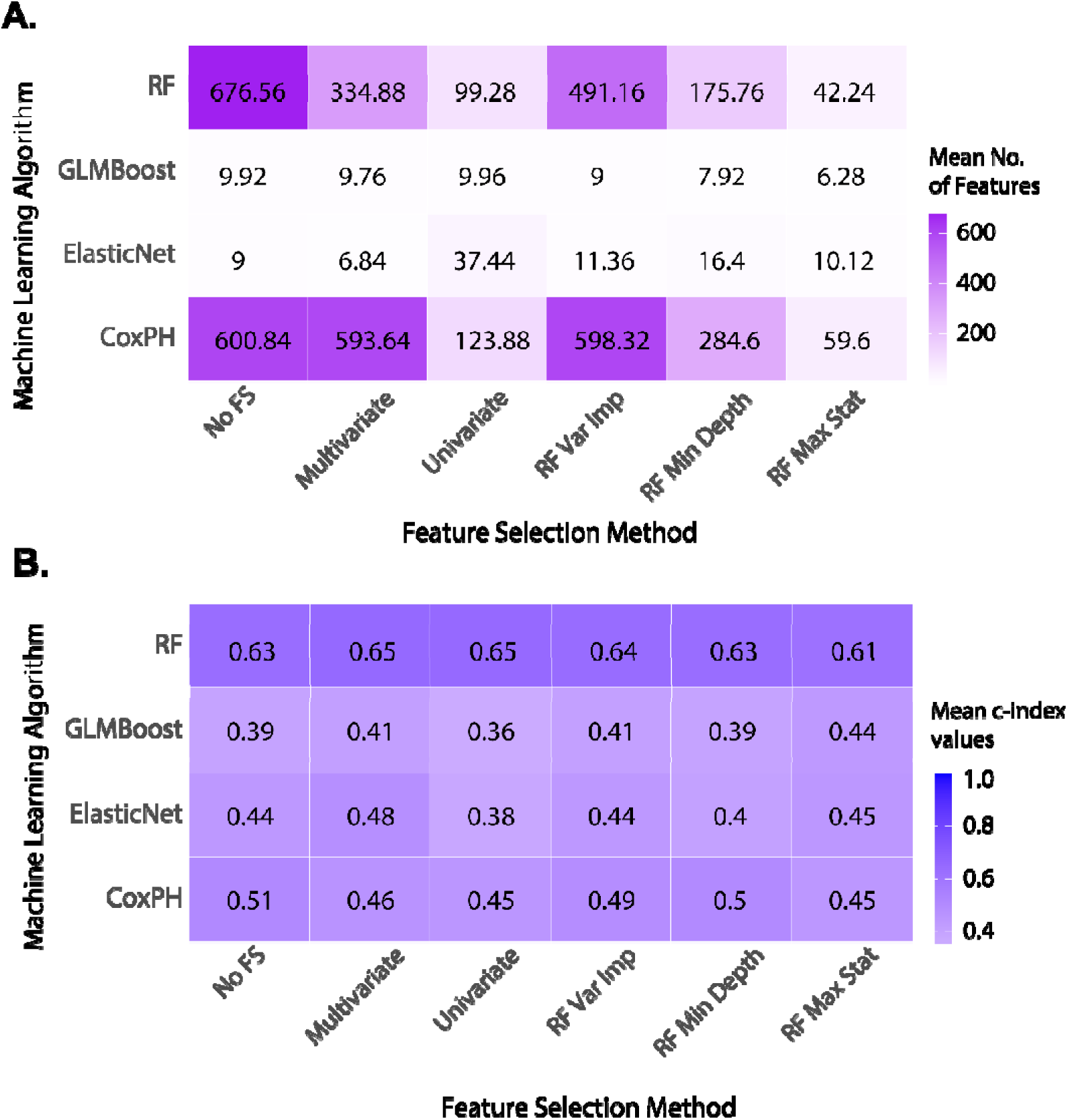
Heatmap of both features selected from each model and filter methods, using BRCA DM data. **A.** Mean number of features selected **B.** Mean c-index values. 95% confidence intervals for all mean values reported in this heatmap are provided in Supplementary Table 6. Abbreviation: RF: Random Forest, No FS: No Feature Selection, Uni/Multivariate: Uni/Multivariate CoxPH, Var Imp: Variable Importance, Min Depth: Minimal Depth, Max Stat: Variable Hunting, GLMBoost: Boosted CoxPH, ElasticNet: Penalised CoxPH

**Table 2.**
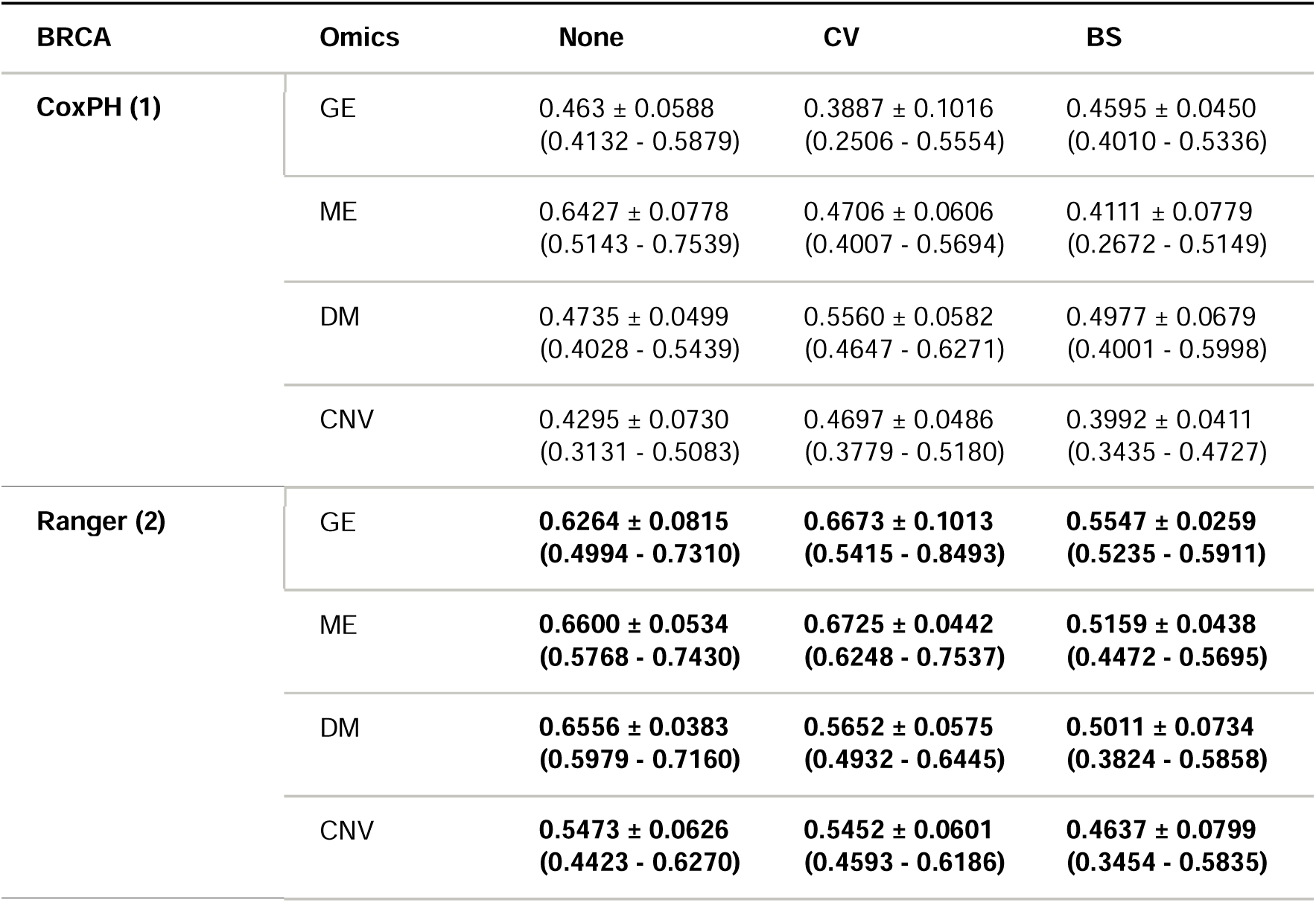
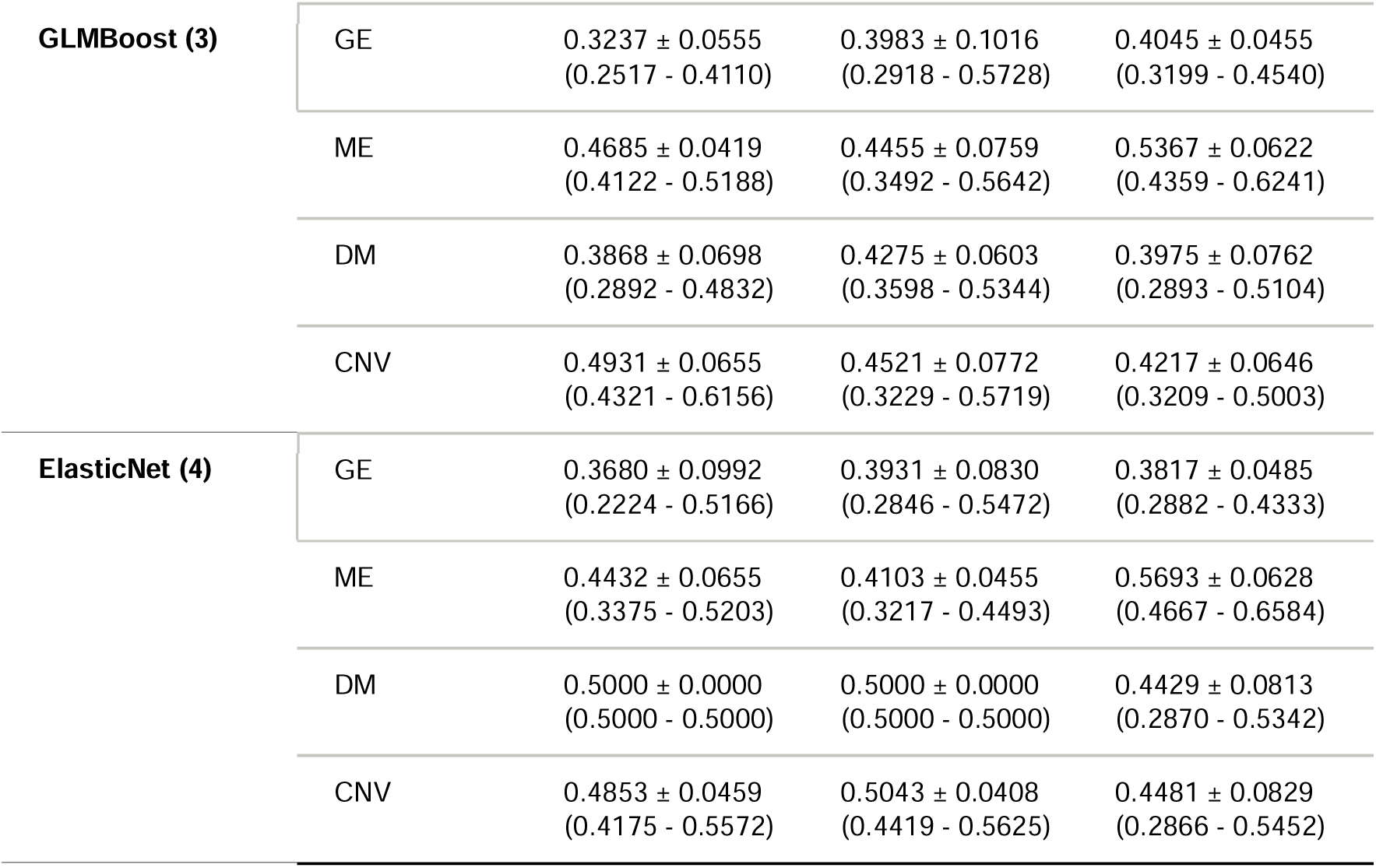
Performance of the feature selection pipeline applied to the BRCA dataset. Results include comparisons to a baseline model (denoted as "None"), where no feature selection is applied. Ranger refers to Random Forest, GLMBoost refers to Boosted Cox, and ElasticNet refers to Penalized Cox. Here the data is represented by the mean cindex with its standard deviation. The numbers in the brackets represent the 95% Confidence Intervals.

| BRCA | Omics | None | CV | BS |
| --- | --- | --- | --- | --- |
| **CoxPH (1)** | GE | 0.463 ± 0.0588<br>(0.4132 - 0.5879) | 0.3887 ± 0.1016<br>(0.2506 - 0.5554) | 0.4595 ± 0.0450<br>(0.4010 - 0.5336) |
|  | ME | 0.6427 ± 0.0778<br>(0.5143 - 0.7539) | 0.4706 ± 0.0606<br>(0.4007 - 0.5694) | 0.4111 ± 0.0779<br>(0.2672 - 0.5149) |
|  | DM | 0.4735 ± 0.0499<br>(0.4028 - 0.5439) | 0.5560 ± 0.0582<br>(0.4647 - 0.6271) | 0.4977 ± 0.0679<br>(0.4001 - 0.5998) |
|  | CNV | 0.4295 ± 0.0730<br>(0.3131 - 0.5083) | 0.4697 ± 0.0486<br>(0.3779 - 0.5180) | 0.3992 ± 0.0411<br>(0.3435 - 0.4727) |
| **Ranger (2)** | GE | **0.6264 ± 0.0815**<br>**(0.4994 - 0.7310)** | **0.6673 ± 0.1013**<br>**(0.5415 - 0.8493)** | **0.5547 ± 0.0259**<br>**(0.5235 - 0.5911)** |
|  | ME | **0.6600 ± 0.0534**<br>**(0.5768 - 0.7430)** | **0.6725 ± 0.0442**<br>**(0.6248 - 0.7537)** | **0.5159 ± 0.0438**<br>**(0.4472 - 0.5695)** |
|  | DM | **0.6556 ± 0.0383**<br>**(0.5979 - 0.7160)** | **0.5652 ± 0.0575**<br>**(0.4932 - 0.6445)** | **0.5011 ± 0.0734**<br>**(0.3824 - 0.5858)** |
|  | CNV | **0.5473 ± 0.0626**<br>**(0.4423 - 0.6270)** | **0.5452 ± 0.0601**<br>**(0.4593 - 0.6186)** | **0.4637 ± 0.0799**<br>**(0.3454 - 0.5835)** |
| **GLMBoost (3)** | GE | 0.3237 ± 0.0555<br>(0.2517 - 0.4110) | 0.3983 ± 0.1016<br>(0.2918 - 0.5728) | 0.4045 ± 0.0455<br>(0.3199 - 0.4540) |
|  | ME | 0.4685 ± 0.0419<br>(0.4122 - 0.5188) | 0.4455 ± 0.0759<br>(0.3492 - 0.5642) | 0.5367 ± 0.0622<br>(0.4359 - 0.6241) |
|  | DM | 0.3868 ± 0.0698<br>(0.2892 - 0.4832) | 0.4275 ± 0.0603<br>(0.3598 - 0.5344) | 0.3975 ± 0.0762<br>(0.2893 - 0.5104) |
|  | CNV | 0.4931 ± 0.0655<br>(0.4321 - 0.6156) | 0.4521 ± 0.0772<br>(0.3229 - 0.5719) | 0.4217 ± 0.0646<br>(0.3209 - 0.5003) |
| **ElasticNet (4)** | GE | 0.3680 ± 0.0992<br>(0.2224 - 0.5166) | 0.3931 ± 0.0830<br>(0.2846 - 0.5472) | 0.3817 ± 0.0485<br>(0.2882 - 0.4333) |
|  | ME | 0.4432 ± 0.0655<br>(0.3375 - 0.5203) | 0.4103 ± 0.0455<br>(0.3217 - 0.4493) | 0.5693 ± 0.0628<br>(0.4667 - 0.6584) |
|  | DM | 0.5000 ± 0.0000<br>(0.5000 - 0.5000) | 0.5000 ± 0.0000<br>(0.5000 - 0.5000) | 0.4429 ± 0.0813<br>(0.2870 - 0.5342) |
|  | CNV | 0.4853 ± 0.0459<br>(0.4175 - 0.5572) | 0.5043 ± 0.0408<br>(0.4419 - 0.5625) | 0.4481 ± 0.0829<br>(0.2866 - 0.5452) |

Before integration, omics datasets are normalized using min-max scaling to standardize values. RFE is then applied iteratively, ranking features based on RF variable importance scores and progressively eliminating the least informative ones. The final multi-omics signature is determined by maximizing predictive performance, measured by the c-index. By refining feature selection both within individual omics and across the integrated dataset, our approach optimizes the signature panel, reducing its size while preserving predictive power. This ensures a more interpretable and clinically applicable model for survival prediction in cancer.

### Pan-cancer Analysis

With the optimal multi-omics combinations identified for BRCA, OV, CESC, and UCEC, we now turn to our pan-cancer analysis to uncover shared multi-modal signatures across these cancers. First, we will conduct an overlap analysis to identify common features across the four cancers and explore potential overlaps by analyzing miRNA targets associated with these features. Next, we will examine disease association networks to understand the relationships between these overlapping targets, offering insights into their interconnected roles in cancer biology. To uncover shared biological themes, we will perform Gene Set Enrichment Analysis (GSEA) on the targets from each cancer, focusing on common Gene Ontology (GO) terms and KEGG pathways. Finally, to ensure the robustness and biological relevance of our findings, we will validate key features using Kaplan-Meier plots and cross-reference them with existing literature. While our feature selection was driven by predictive power in survival analysis, this validation step will confirm that these features have meaningful biological implications. Through this integrated approach, we aim to demonstrate that our results are not only statistically significant but also provide valuable biological insights into the commonalities and differences across the cancers under study.

## Results

All analyses were conducted on the University of New South Wales (UNSW) Katana high-performance computing platform, utilizing 16 CPUs and 124 GB of memory per job to efficiently process large-scale omics data. While optimized for HPC, the pipeline can also run on local machines by reducing data size or enabling parallelization for improved runtime efficiency.

### Evaluation of Individual Filter Methods

Before developing our feature selection pipeline, we evaluated the four filter methods used in this study (Figure 1). Due to computational constraints, we limited our analysis to these four methods. The results are presented as heatmaps showing the mean C-index values and the number of features selected across 5 repeats of 5-fold cross-validation for each combination of ML algorithms (rows) and filter methods (columns). The CPH model, included as a benchmark, is compared with the other models. Column 1, representing results without feature selection, serves as the baseline for comparison.

Figure 4 presents the results from the BRCA DM dataset, which reflect trends consistent across other modalities and cancer types (Supplementary Tables 6 to 9), serving as a representative example. We expected the performance of each filter method to at least match the baseline (no feature selection). As shown in (A), feature selection resulted in a significant reduction in the number of features, with predictive performance remaining stable or slightly decreasing, indicating the removal of redundant features. The RF model consistently outperformed others, with the lowest C-index being 0.61. Surprisingly, both Penalized and Boosted Cox models underperformed compared to the baseline CPH, with C-index values dropping below 0.5. This was unexpected, given their design to handle high-dimensional data. However, applying filter methods did generally improve the performance of these extended Cox models. Based on overall performance, we decided to remove the univariate filter method, as it evaluates features in isolation, limiting its ability to capture the combined effect of features.

### Performance of Feature Selection Pipelines

In this study, we developed two feature selection pipelines: one based on cross-validation and the other on bootstrapping, both utilizing our four main filter methods. Table 2 compares the performance of these pipelines to a baseline with no feature selection. Column 1 lists the predictive models, while column 2 presents the modalities evaluated. Overall, the cross-validation pipeline showed promising results, with improvements in some areas and slight reductions in others. In contrast, the bootstrapping pipeline required significantly more computational resources and yielded poorer performance overall. As with our filter method performance test, Random Forest consistently delivered the best results. Based on the BRCA dataset, we decided to adopt CV as our primary feature selection pipeline moving forward.

Supplementary Tables 10 to 12 show the results for the other cancers investigated using the CV pipeline. Overall, Random Forest appears to be the highest-performing predictive model across all four cancers. OV has an overall lower c-index compared to the other cancers.

### One-Stage vs. Two-Stage Refinement

For our multi-omics integration, we applied two techniques: One-Stage and Two Refinement, as seen in Figures 3.

In Table 3, we illustrate the performance comparison between One-Stage and Two-Stage Refinement using the CESC dataset. This trend is consistent across all cancer types tested, not just CESC (*Supplementary Tables 14 to 16*). Our results show that Two-Stage Refinement consistently outperforms One-Stage Refinement. For example, in the CESC dataset, One-Stage Refinement achieved a c-index of 0.70, while Two-Stage Refinement significantly improved this to 0.80. This suggests that the optimal modality combination can vary depending on the integration strategy used. In this case, Two-Stage Refinement identified the combination of DM, ME, and CNV as superior to the DM and ME combination preferred by One-Stage Refinement. Overall, Two-Stage Refinement demonstrated better performance across all cancer datasets, leading us to select it as our primary integration strategy.

**Table 3.**
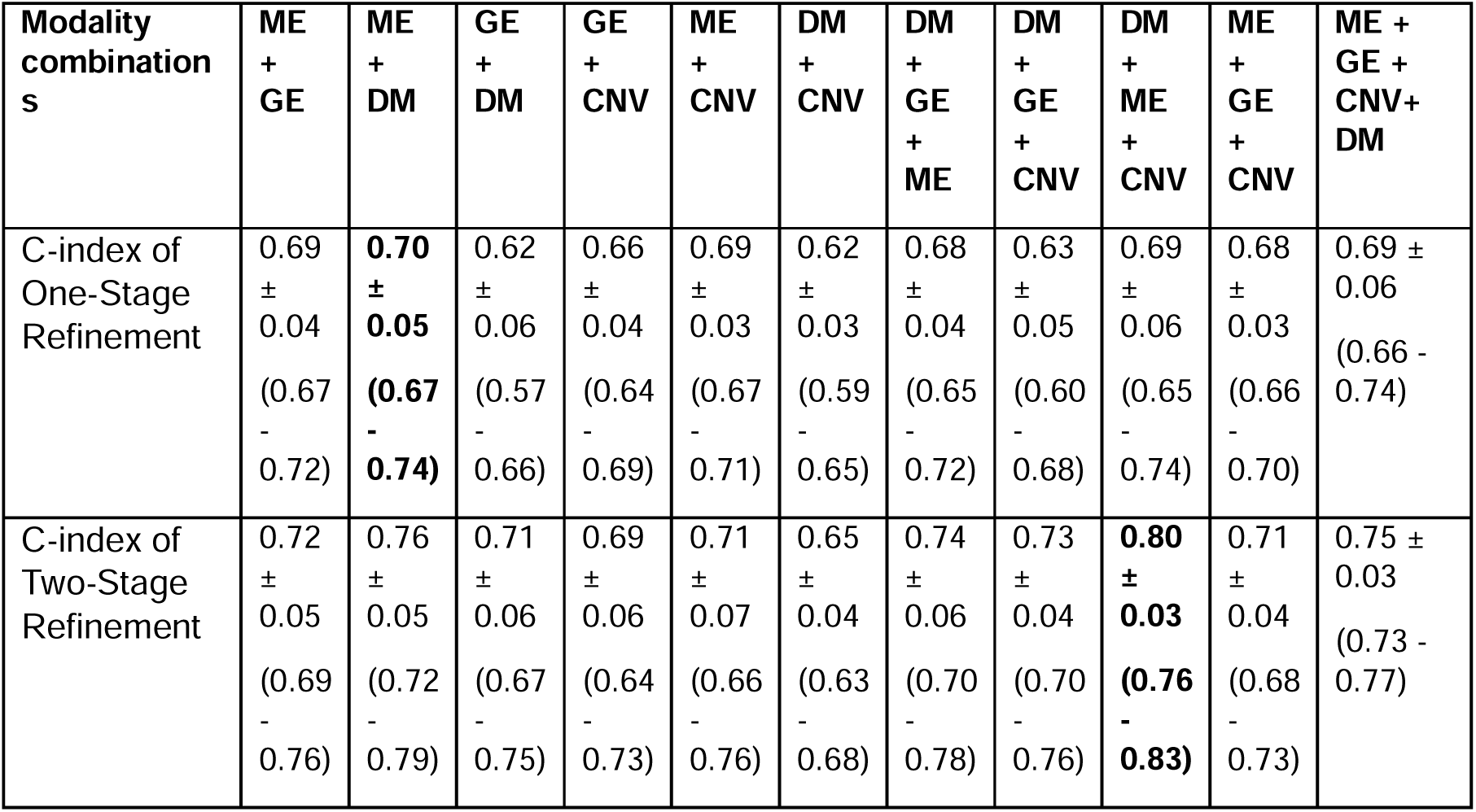
Comparison between One-Stage and Two-Stage Refinement using CESC dataset.

### Multi-omics results

Using Two-Stage Refinement as our main integration method, we tested every possible modality combination and observed their c-index.

Table 4 presents the results for each modality combination across all cancers. For BRCA, the highest performance was achieved with a c-index of 0.77 using a combination of DNA methylation, miRNA expression, and copy number variation. This combination outperformed the baseline single-modality Random Forest model, which achieved a maximum c-index of 0.6725 with gene expression data alone. *Supplementary Tables 17 to 20* details the features from each modality that contributed to this peak performance. For OV, which showed the lowest performance relative to the other cancers, the best-performing modality combination was miRNA expression, gene expression, and DNA methylation, achieving a c-index of 0.65. This combination outperformed the single-omics models, although it had fewer contributing features compared to the other cancers. Like BRCA, the combination of DNA methylation, miRNA expression, and copy number variation yielded the highest performance for CESC, achieving a c-index of 0.80, with miRNA expression being the most influential. For UCEC, the same combination achieved a c-index of 0.76, with miRNA expression as the major contributor. Notably, this modality combination was the most predictive for three out of four cancer types, and DNA methylation and miRNA expression were consistently present in the final integration for all cancers.

**Table 4.**
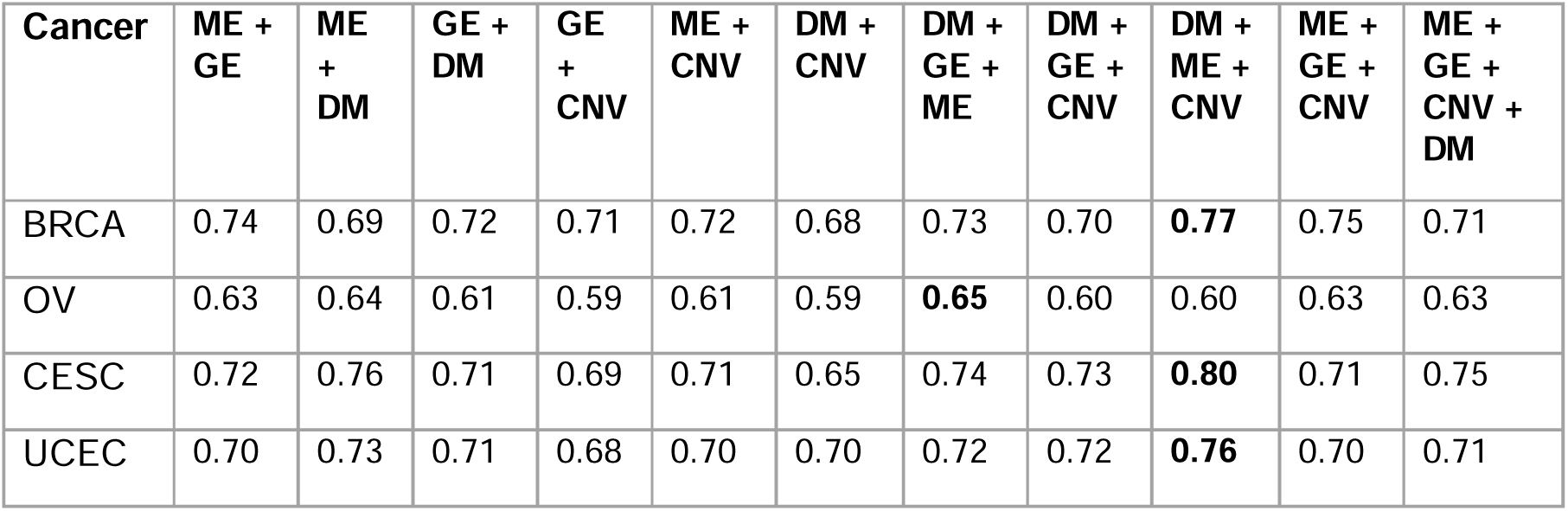
Multi-omics results for all cancers using Two-Stage Refinement.

| Cancer | ME +<br>GE | ME<br>+<br>DM | GE +<br>DM | GE<br>+<br>CNV | ME +<br>CNV | DM +<br>CNV | DM +<br>GE +<br>ME | DM +<br>GE +<br>CNV | DM +<br>ME +<br>CNV | ME +<br>GE +<br>CNV | ME +<br>GE +<br>CNV +<br>DM |
| --- | --- | --- | --- | --- | --- | --- | --- | --- | --- | --- | --- |
| BRCA | 0.74 | 0.69 | 0.72 | 0.71 | 0.72 | 0.68 | 0.73 | 0.70 | **0.77** | 0.75 | 0.71 |
| OV | 0.63 | 0.64 | 0.61 | 0.59 | 0.61 | 0.59 | **0.65** | 0.60 | 0.60 | 0.63 | 0.63 |
| CESC | 0.72 | 0.76 | 0.71 | 0.69 | 0.71 | 0.65 | 0.74 | 0.73 | **0.80** | 0.71 | 0.75 |
| UCEC | 0.70 | 0.73 | 0.71 | 0.68 | 0.70 | 0.70 | 0.72 | 0.72 | **0.76** | 0.70 | 0.71 |

### Pan-cancer Analysis - Overlapping Features

After obtaining the final set of multi-modal features for each cancer using our multi-omics pipeline, we conducted a pan-cancer analysis to identify overlapping features among cancers.

Table 5 presents the results of our direct overlap analysis, highlighting the shared pan-cancer signatures among the cancers. The most overlapping features were observed between BRCA and UCEC, with methylation signatures comprising many of the overlapping features, and between CESC and UCEC, where miRNA signatures predominated. No feature appeared in all four cancers, with only the miRNA signature mir.150 being common to three cancers: BRCA, CESC, and UCEC.

**Table 5.**
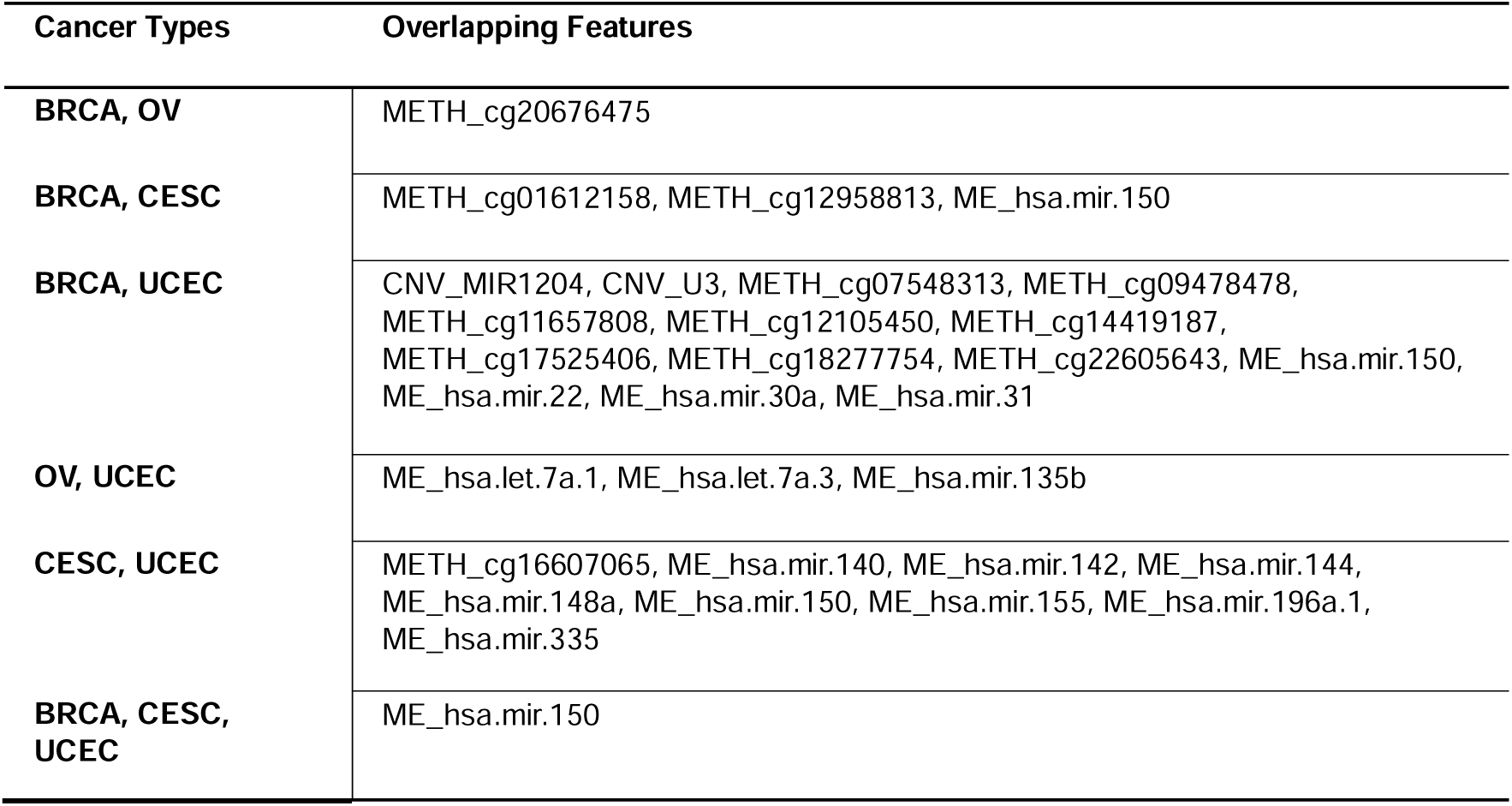
Directly overlapping features across all cancers, METH = Methylation, CNV = Copy Number Variation, ME = miRNA.

### MiRNA Features and Targets

The goal of this analysis is to uncover further overlaps among our cancers beyond the direct feature overlap comparison. This involves examining the miRNA targets of our miRNA features from each cancer to identify overlapping gene targets. Using the multiMiR package, we extracted validated miRNA-target interactions and filtered these validated gene targets to include only those expressed in our studied cancers (Figure 5).

**Figure 5.**
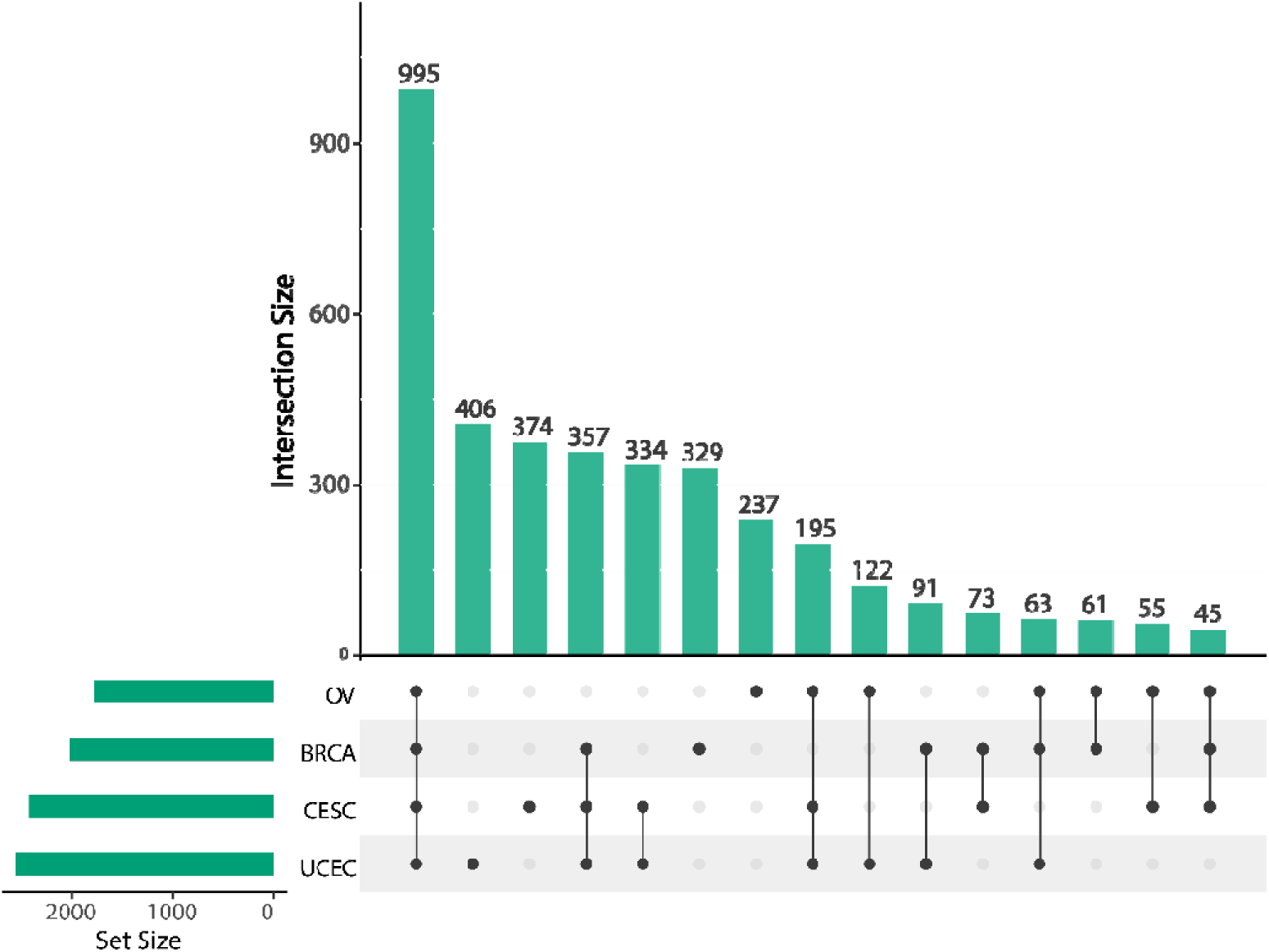
Upset plot showing direct overlapping miRNA Targets between cancers

We identified that majority of the gene targets are shared across all four cancers, with 995 targets. Notably, UCEC contributed the most gene targets, while OV provided the least. Using the 995 overlapping gene targets, we performed enrichment analysis using DisGeNET, a database of gene-disease associations.

Figure 6(A) presents an enrichment map that highlights a large, interconnected disease cluster, where genes with shared associations group together into functionally related modules. Within this overarching cluster, we observe that multiple adenocarcinomas— including those of the colon, esophagus, and stomach—are closely linked. Likewise, various carcinomas, such as breast, intra- and ductal, and gallbladder carcinomas, integrate into the same network. Additionally, meningiomas and fibrosarcomas are embedded within this cluster, suggesting shared molecular pathways among these malignancies. These findings highlight extensive gene overlap across different cancers and related conditions, reinforcing the idea of common biological mechanisms underlying their development.

**Figure 6.**
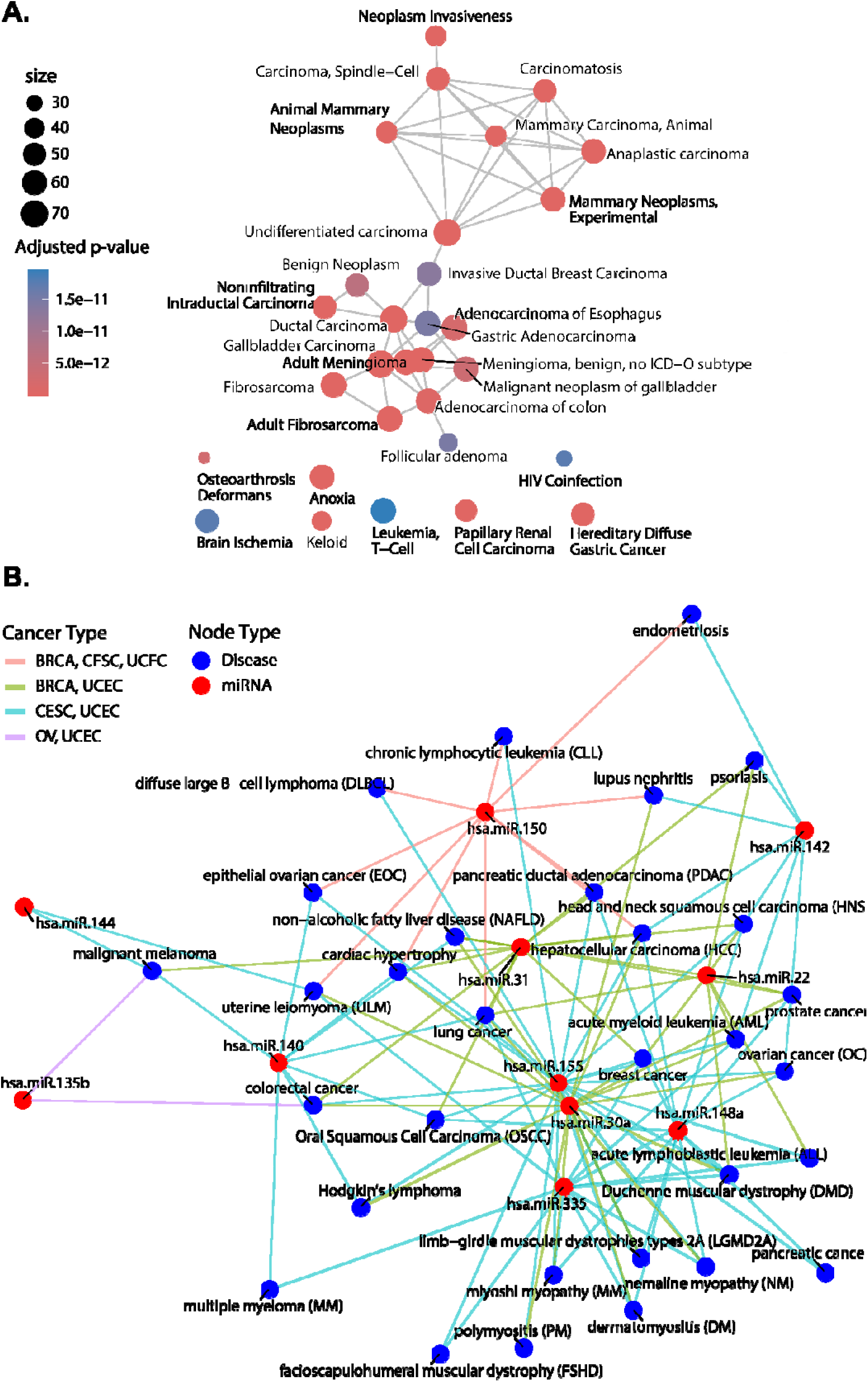
Disease network using the overlapping targets and miRNA. **(A)** Enrichment Map of overlapping miRNA Targets disease network. **(B)** Overlapping miRNA Disease associations across cancers.

Figure 6(B) focuses on the overlapping miRNA features identified in our direct overlap analysis (Table 8)—those miRNA signatures present in two or more cancers. To further validate their biological relevance, we explored the miRNAs’ disease associations using the mir2disease database. Most of the associated disease terms are linked to various cancers, such as carcinoma, lymphoma, leukemia, myeloma, and melanoma, suggesting that these miRNAs may play roles in multiple oncogenic processes and molecular pathways. The recurrence of these cancer types in the disease association networks indicates that these miRNAs could regulate genes and pathways critical for tumorigenesis and cancer progression across different cancer types, pointing to potential shared oncogenic mechanisms and common therapeutic targets. Additionally, the presence of muscle-related diseases, like muscular dystrophy and polymyositis, in the network suggests that these miRNAs might also influence muscle-related pathways, highlighting potential shared biological mechanisms between cancer and muscle diseases, as supported by previous studies [25].

### Gene Set Enrichment Analysis (GSEA)

We performed Gene Set Enrichment Analysis (GSEA) on our miRNA targets to identify shared biological themes and validate our signatures across cancers.

Figure 7(A) shows the enrichment plot for BRCA miRNA targets, highlighting GO terms like cytoplasmic translation, biosynthetic processes, and protein localization regulation, which are critical for cancer proliferation. Overexpression of translation initiation factors (e.g., eIF4E) and dysregulated telomerase activity are linked to poor prognosis in breast cancer [26,27]. MiRNAs involved in RNA binding and processing play key roles in post-transcriptional regulation. The KEGG plot identifies pathways like protein processing in the ER and proteoglycans in cancer, which impact cell adhesion, signaling, and metastasis [28]. Additionally, pathways related to infectious diseases and neurodegenerative disorders, such as Alzheimer’s and Huntington’s diseases, intersect with cancer biology [29]. Figure 7(B) presents enrichment results for OV miRNA targets, where GO terms emphasize protein synthesis, and KEGG pathways align with BRCA results, highlighting cancer-related and signaling pathways. Figure 7(C) shows enrichment for CESC miRNA targets, with the Interleukin-27 (IL-27)-Mediated Signaling Pathway emerging as key in metastasis suppression and tumor migration [30]. KEGG pathways reflect trends in essential cellular processes and cancer-related pathways. Figure 7(D) presents results for UCEC miRNA targets. GO terms focus on the regulation of apoptotic signaling by p53, a critical pathway often disrupted in cancer through TP53 mutations, allowing cells to evade apoptosis [31]. KEGG pathways highlight fundamental cellular processes and cancer-related and disease-associated pathways.

**Figure 7.**
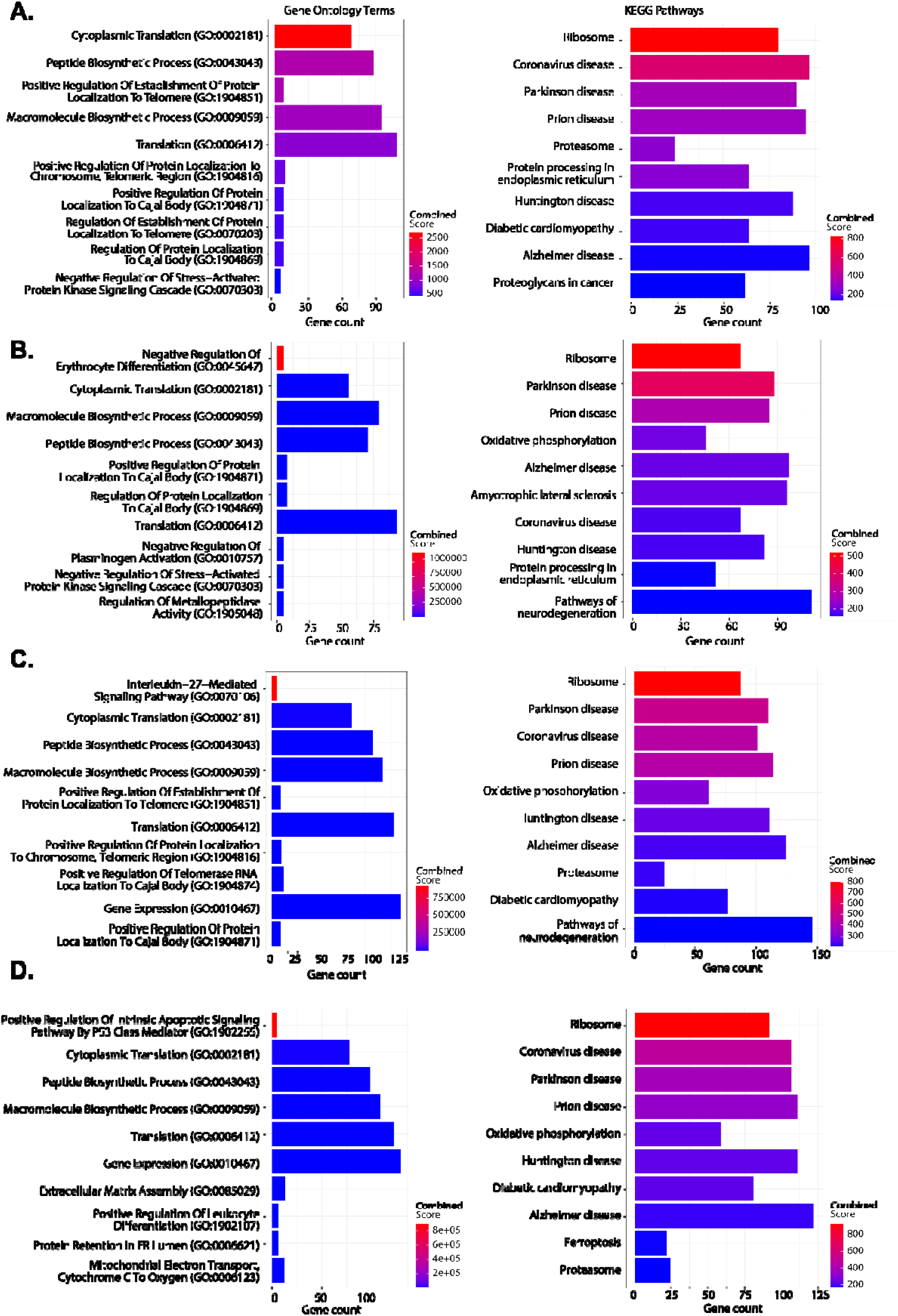
GSEA enrichment plots of Top 10 GO Terms and KEGG Pathways. **(A)** BRCA, **(B)** OV, **(C)** CSEC and **(D)** UCEC

### Analyses of the Overlap of Enriched Functions

After reviewing the GSEA results for each cancer’s miRNA targets and identifying notable trends, we analyzed the overlapping GO Terms and KEGG Pathways across the cancers to identify shared biological themes and pathways, further elucidating underlying mechanisms and potential therapeutic targets.

Figure 8(A) shows an upset plot of GO Term and KEGG Pathway overlaps, revealing the largest intersection when all four cancers (BRCA, OV, UCEC, and CESC) share common GO terms. This suggests shared biological processes regulated by the miRNAs across these cancer types, indicating potential common pathways for therapeutic exploration.

**Figure 8.**
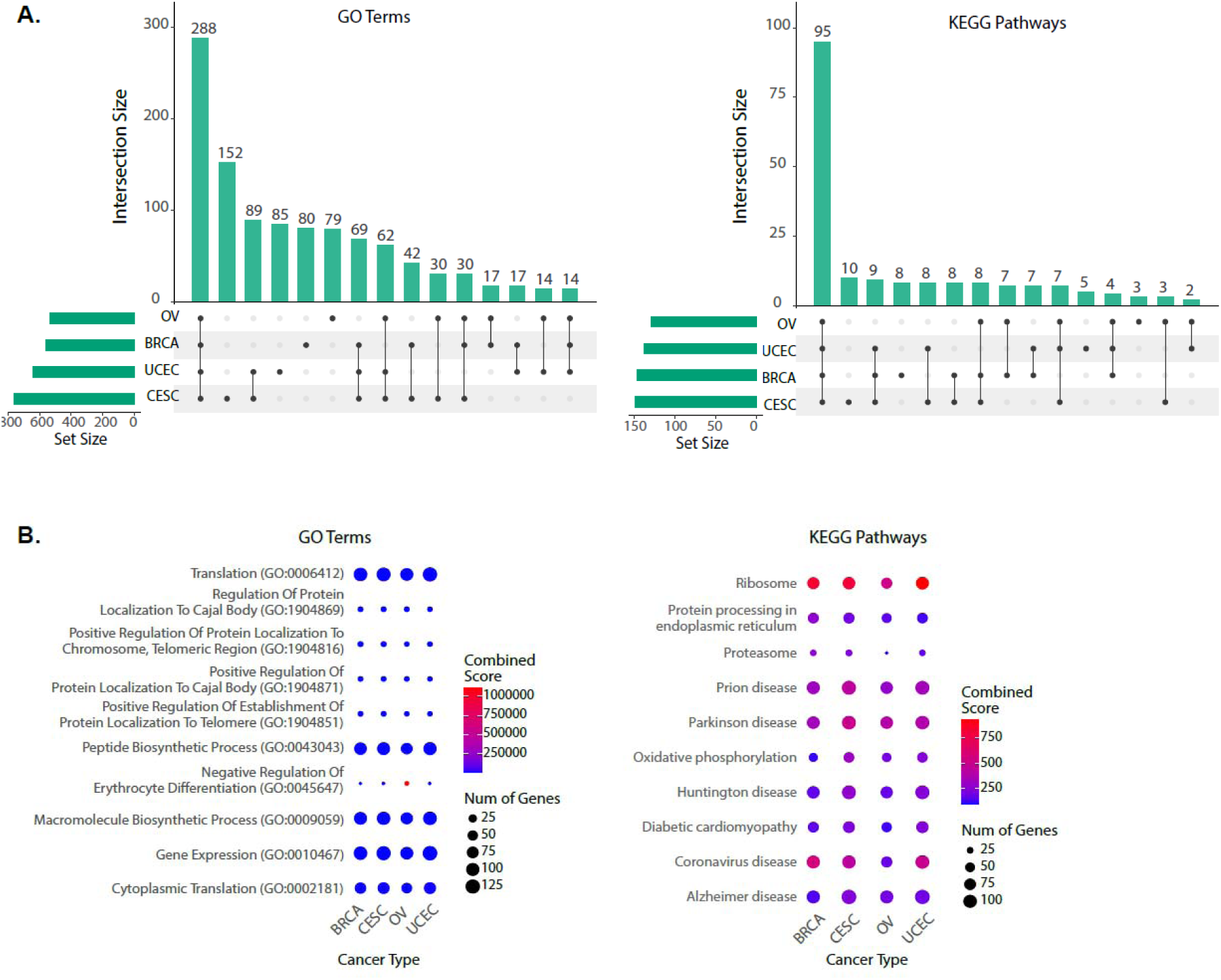
**A.** Upset plot showing GSEA overlaps across all cancers for miRNA targets. B. Enrichment dot plot showing Top 10 GSEA overlaps across all cancers.

Figure 8(B) presents the top 10 shared GO Terms and KEGG pathways. The GO terms highlight key processes like cell growth, protein synthesis, and RNA regulation, which are frequently dysregulated in cancer and contribute to hallmarks such as uncontrolled proliferation and resistance to cell death [26–31]. Shared pathways like "Ribosome" and "Protein processing in the endoplasmic reticulum" emphasize protein synthesis [32], while infectious disease and neurodegenerative disease pathways underscore broader clinical relevance. These results not only confirm the miRNAs’ role in cancer but also their potential impact on other health conditions.

### Kaplan-Meier Survival Analysis and Significance Testing

To validate the biological relevance of our overlapping multi-modal signatures, we conducted Kaplan-Meier (KM) survival analyses and cross-referenced the results with existing literature, confirming their potential impact on cancer prognosis. First, we explored miR-22, present in both UCEC and BRCA (Table 5). Koufaris et al. [33] showed that in BRCA, miR-22 represses glycolytic metabolism, reducing survival outcomes, acting as an oncomir. In contrast, Cui et al. [34] found miR-22 inhibits EMT in liver cancer, acting as a tumor suppressor. Figure 9(A) supports these findings, showing that high miR-22 levels are associated with lower survival in BRCA, while low levels in UCEC are linked to worse survival, suggesting a tumor suppressor role in UCEC. Next, we examined the cg probe cg17525406, hypermethylated in lung cancer and linked to tumor suppressor gene silencing [35,36]. In BRCA, high methylation of cg17525406 correlates with lower survival (Figure 9(B)), confirming its role in silencing tumor suppressors. However, the opposite is observed in UCEC, where low methylation correlates with lower survival, suggesting a dual role of cg17525406 as both an oncogene and tumor suppressor depending on the cancer type. Lastly, we explored miR-150, which appears in at least three cancers. Wang, Ren, and Zhang [37], Sugita et al. [38], and Sun et al. [39] all reported deregulation of miR-150 in cancers. Figure 9(C) shows that low miR-150 expression in UCEC, BRCA, and CESC correlates with reduced survival, supporting its role as a potential cancer prognosis biomarker and therapeutic target.

**Figure 9.**
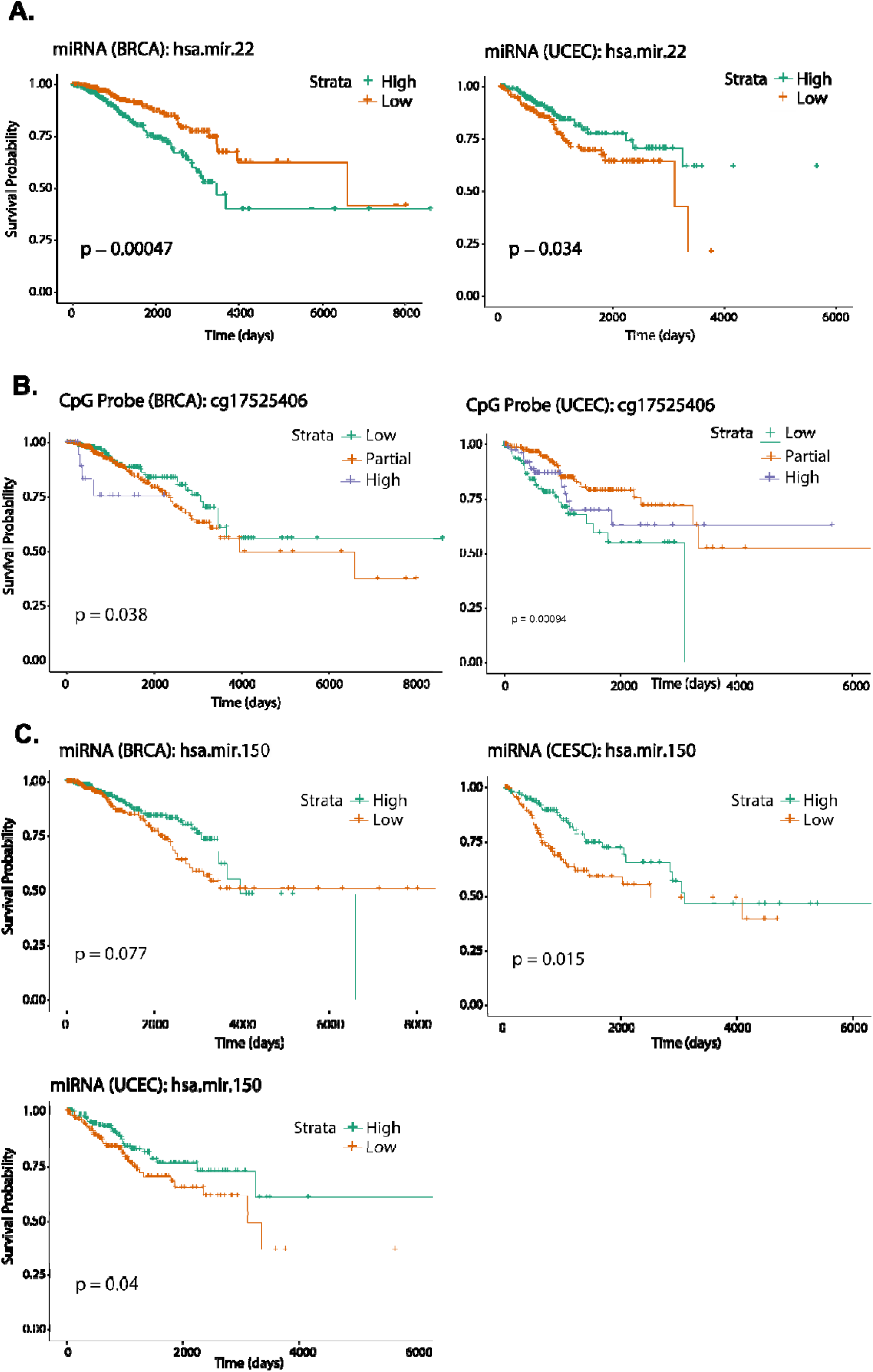
Kaplan-Meier Survival plots **(A)** miR-22, **(B)** cg17525406, **(C)** miR-150

## Discussion

PRISM was developed to (1) improve cancer survival prediction using multi-omics data and feature selection methods and (2) identify key multi-modal features influencing survival outcomes. It provides a comprehensive framework for feature selection and survival modeling, advancing prognostic marker discovery. By benchmarking multiple feature selection methods alongside survival models, PRISM ensures robust performance evaluation. Its statistically rigorous strategy integrates cross-validation with ensemble voting to enhance reproducibility and resilience to patient cohort variations. PRISM optimizes prognostic signatures through recursive feature elimination, reducing redundancy while maintaining predictive accuracy, improving clinical feasibility and cost-effectiveness. It integrates multi-omics data through a two-stage refinement, selecting features within individual omics layers before optimizing at both single- and multi-omics levels. As a fully integrated pipeline, PRISM streamlines the entire workflow, from data retrieval and preprocessing to survival modeling and functional analysis, ensuring broad applicability beyond TCGA datasets.

We drew inspiration from previous studies, including Zhao et al. [1], who identified pan-cancer prognostic biomarkers through multi-omics integration, and Tong et al. [19], who explored deep learning-based feature integration for breast cancer survival. Drawing from Spooner et al. [3] for machine learning models and feature selection, we demonstrated that multi-modal integration enhances survival predictions over single-omics data. Among the algorithms evaluated, Random Forest was highly effective, particularly for high-dimensional data. Our analysis demonstrated that the optimal multi-modal combination—DM, ME, and CNV—achieved a c-index of 0.77 in BRCA, outperforming Tong et al.’s c-index of 0.641, which utilized only DM and ME data. This highlights the complementary role of CNV in enhancing the predictive power when combined with DM and ME data. Even without the best-performing combination, our basic multi-omics strategy of DM and ME yielded a c-index of 0.69, outperforming Tong et al.’s refined method. This highlights the importance of selecting the right survival model and features before integrating multi-omics data. In our pan-cancer analysis, miR-150 was present in three cancers, and gene target overlaps revealed significant associations with common oncogenic pathways, as well as pathways related to infectious and neurodegenerative disorders. Kaplan-Meier analysis confirmed the biological relevance of these multi-modal signatures in survival outcomes, with some features acting as oncogenes or tumor suppressors depending on the cancer type. These results emphasize the potential of multi-modal integration in identifying critical biomarkers and pathways, offering insights for future cancer therapies.

However, several areas need improvement before expanding the study. First, while we employed pre-processing techniques similar to those of Zhao et al. [1] and Tong et al. [19], the uniform approach across all cancers may have been too broad, particularly for the OV dataset, which showed a lower c-index. Tailoring pre-processing strategies to specific cancer types could improve results. Additionally, we used basic imputation methods, such as mean imputation, whereas more advanced techniques like Multiple Imputation by Chained Equations (MICE) could enhance data quality and model performance. Our CNV data pre-processing was basic, relying on direct Gene Level Scores from TCGA, while more refined methods like GISTIC2.0 from Firebrowse could improve accuracy. Lastly, we did not account for differentially expressed genes (DEGs) and differentially methylated regions (DMRs), which could have enriched the gene features and cg probes. Addressing these limitations will be essential for refining data quality and improving future analyses.

An important consideration for future work is the potential integration of deep learning methods to enhance multi-omics data analysis. However, PRISM aims to identify a minimal yet highly predictive multi-omics signature panel to improve clinical utility and commercial viability. To achieve this, we implemented a rigorous feature selection strategy that combines machine learning-based selection with recursive feature elimination, optimizing panel size while maintaining competitive predictive performance. In contrast, deep learning models, such as autoencoders and transformer-based models, learn features directly from high-dimensional data without explicit feature selection, often resulting in less interpretable features. While prior studies (e.g., References 2, 10, 17–19) successfully applied deep neural networks to multi-omics data, they require the inclusion of all omics features, which can be economically unfeasible due to the high costs of high-throughput omics technologies. Notably, PRISM’s performance on TCGA data is comparable to, and in some cases exceeds, deep learning models. For example, Chai et al. [2] reported test C-index values of 0.66 for BRCA and 0.69 for CESC, while PRISM achieved 0.77 for BRCA and 0.80 for CESC on holdout test data. While direct comparisons are limited, these results emphasize the competitive advantage of PRISM. Nevertheless, we acknowledge the future potential of deep learning methods for multi-omics analysis as computational and economic barriers evolve.

## Data Availability

All datasets used in this study are publicly accessible through The Cancer Genome Atlas (TCGA) data portal. The PRISM pipeline, along with its documentation, is available at the following repository: https://github.com/VafaeeLab/PRISM/.

https://www.cancer.gov/tcga

https://github.com/VafaeeLab/PRISM/.

## Supplementary Table Legends

**Supplementary Table 1.** Comparison of the BRCA dataset, showing the number of features and samples across all modalities before and after the preprocessing steps.

**Supplementary Table 2.** Comparison of preprocessing steps for OV dataset

**Supplementary Table 3.** Comparison of preprocessing steps for CESC dataset

**Supplementary Table 4.** Comparison of preprocessing steps for UCEC dataset

**Supplementary Table 5.** Comprehensive summary of the methods applied in our study, including each method’s key hyperparameters and the corresponding R package utilized for implementation.

**Supplementary Table 6.** Mean performance with standard deviation and Confidence Intervals from each model and filter methods, using BRCA ME data.

**Supplementary Table 7.** Mean performance with standard deviation and Confidence Intervals from each model and filter methods, using BRCA DM data.

**Supplementary Table 8.** Mean performance with standard deviation and Confidence Intervals from each model and filter methods, using BRCA GE data.

**Supplementary Table 9.** Mean performance with standard deviation and Confidence Intervals from each model and filter methods, using BRCA CNV data.

**Supplementary Table 10.** Overview of the performance of the feature selection pipeline applied to the OV dataset.

**Supplementary Table 11.** Overview of the performance of the feature selection pipeline applied to the CESC dataset.

**Supplementary Table 12.** Overview of the performance of the feature selection pipeline applied to the UCEC dataset.

**Supplementary Table 13.** Comparison between One-Stage and Two Refinement using BRCA dataset.

**Supplementary Table 14.** Comparison between One-Stage and Two Refinement using OV dataset.

**Supplementary Table 15.** Comparison between One-Stage and Two Refinement using UCEC dataset.

**Supplementary Table 16.** Comparison between One-Stage and Two Refinement using CESC dataset.

**Supplementary Table 17.** Multi-omics results for BRCA

**Supplementary Table 18.** Multi-omics results for OV

**Supplementary Table 19.** Multi-omics results for CESC

**Supplementary Table 20.** Multi-omics results for UCEC

## Supplementary Figures Legends

**Supplementary Figure 1.** Heatmap of both features selected from each model and filter methods, using BRCA GE data. Mean number of features selected (A), Mean c-index values (B)

**Supplementary Figure 2.** Heatmap of both features selected from each model and filter methods, using BRCA ME data. Mean number of features selected (A), Mean c-index values (B)

**Supplementary Figure 3.** Heatmap of both features selected from each model and filter methods, using BRCA CNV data. Mean number of features selected (A), Mean c-index values (B)

**Supplementary Figure 4:** Distribution of both mir-22 and mir-150 expression across these three age groups.

**Supplementary Figure 5:** KM plot showing survival using BRCA miRNA data stratified by age groups.

## Acknowledgements

The results published here are in whole or part based upon data generated by the TCGA Research Network: https://www.cancer.gov/tcga. This research includes computations using the computational cluster Katana, supported by Research Technology Services at UNSW Sydney (https://research.unsw.edu.au/research-technology-services-restech).

## Author Contribution

Conceptualization, F.V.; Methodology, F.V. and R.N.; Software, F.V. and R.N.; Formal Analysis, F.V. and R.N.; Investigation, F.V.; Data Curation, R.N.; Writing – Original Draft Preparation, F.V. and R.N.; Writing – Review & Editing, F.V. and R.N.; Visualization, F.V. and R.N.; Supervision, F.V.; Project Administration, F.V.; Funding Acquisition, Not Applicable.

## Declaration

This study relies on the use of publicly accessible data from The Cancer Genome Atlas (TCGA) data and does not involve human participants directly. The study complies with the respective data access policies and data use certification agreement as outlined by TCGA Ethics and Policies (https://www.cancer.gov/ccg/research/genome-sequencing/tcga/history/ethics-policies).

We also declare that during the preparation of this work, the authors utilized GPT-4 to enhance the clarity and readability of the content in some sections. All content was originally generated by the authors. Following the use of this tool/service, the authors thoroughly reviewed and edited the material as necessary and take full responsibility for the final content of the publication.

