## Supplementary Figures for "Multi-Omics Prognostic Marker Discovery and Survival Modeling: A Case Study on Pan-Cancer Survival Analyses in Women’s Cancers"

**List of Supplementary Figures:**

**Supplementary Figure 1.** Heatmap of both features selected from each model and filter methods, using BRCA GE data. Mean number of features selected (A), Mean c-index values (B)

**Supplementary Figure 2.** Heatmap of both features selected from each model and filter methods, using BRCA ME data. Mean number of features selected (A), Mean c-index values (B)

**Supplementary Figure 3.** Heatmap of both features selected from each model and filter methods, using BRCA CNV data. Mean number of features selected (A), Mean c-index values (B)

**Supplementary Figure 4:** Distribution of both mir-22 and mir-150 expression across these three age groups.

**Supplementary Figure 5:** KM plot showing survival using BRCA miRNA data stratified by age groups.

**Supplementary Figure 1.** Heatmap of both features selected from each model and filter methods, using BRCA GE data. Mean number of features selected (A), Mean c-index values (B)

**A.**

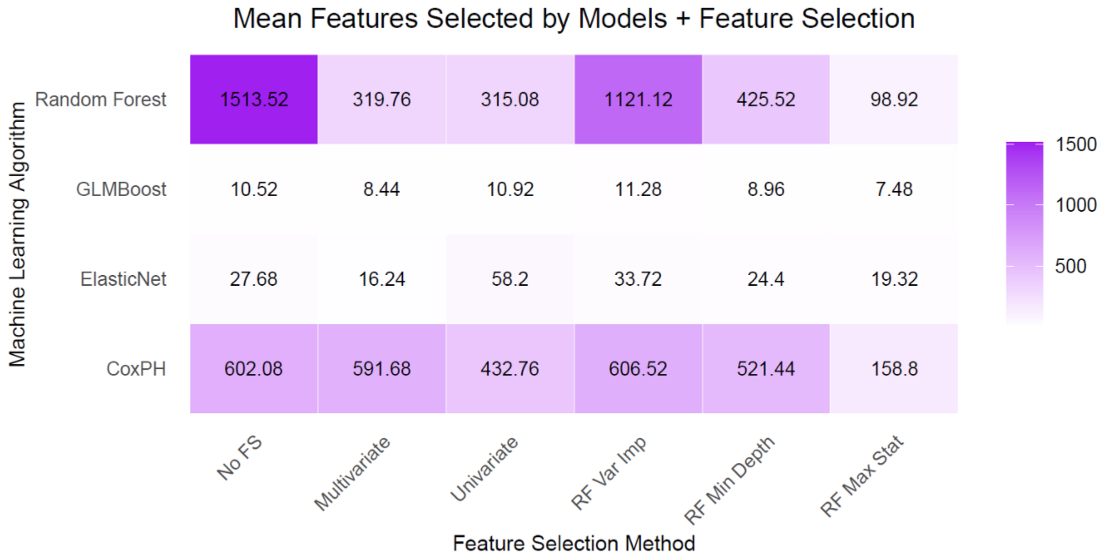


**B.**

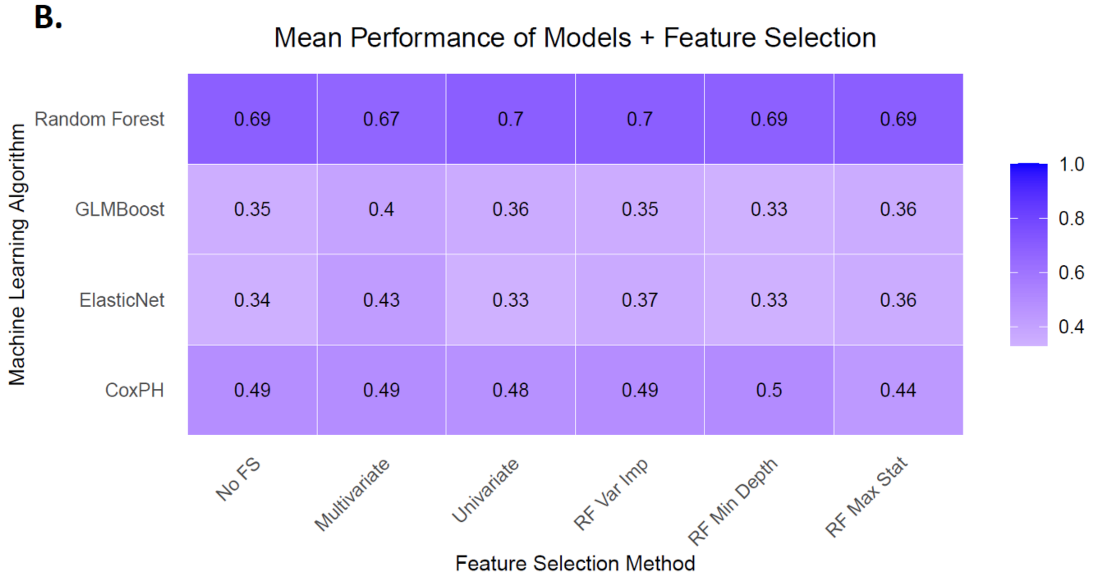

**Supplementary Figure 2.** Heatmap of both features selected from each model and filter methods, using BRCA ME data. Mean number of features selected (A), Mean c-index values (B)

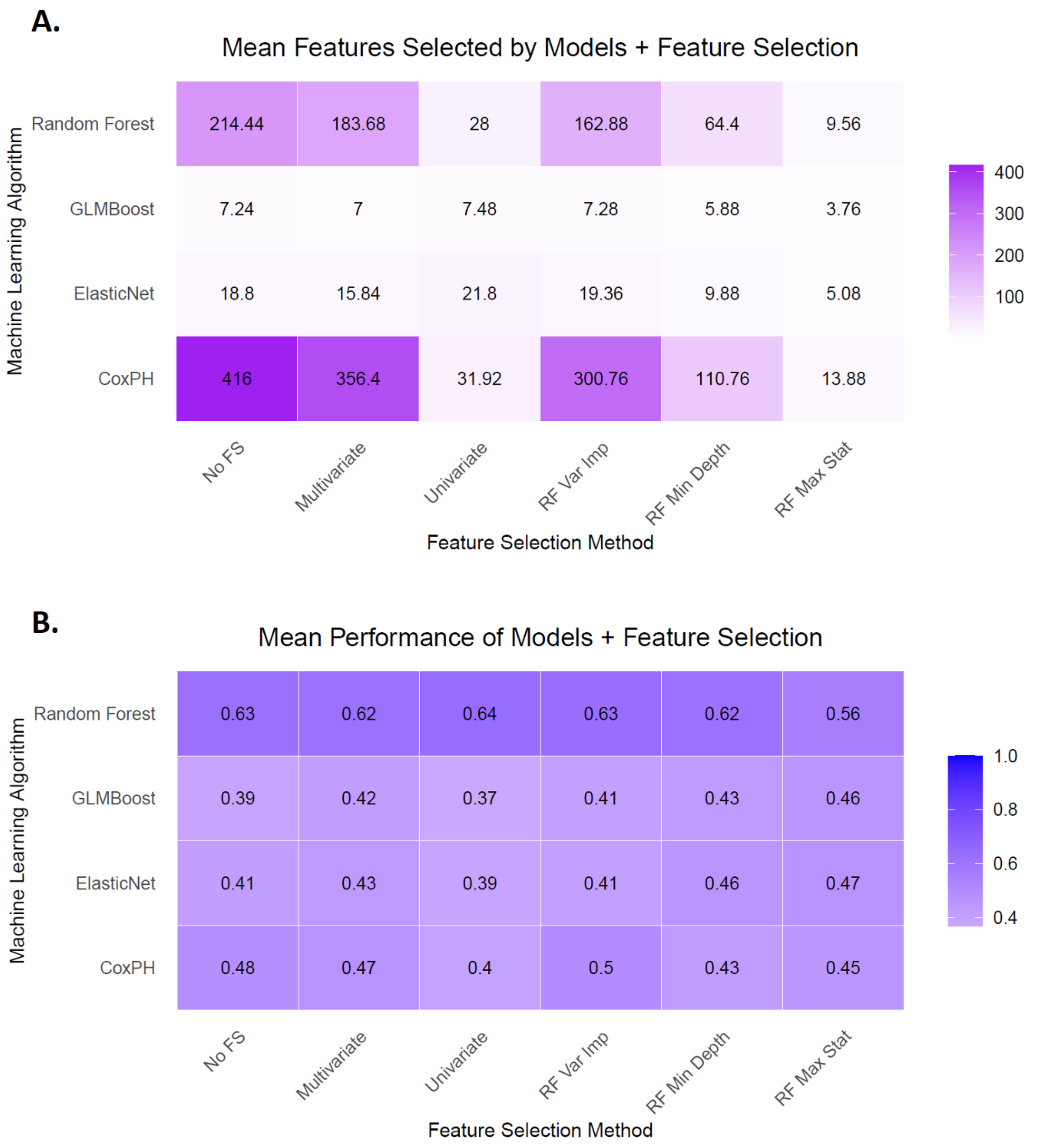

**Supplementary Figure 3.** Heatmap of both features selected from each model and filter methods, using BRCA CNV data. Mean number of features selected (A), Mean c-index values (B)

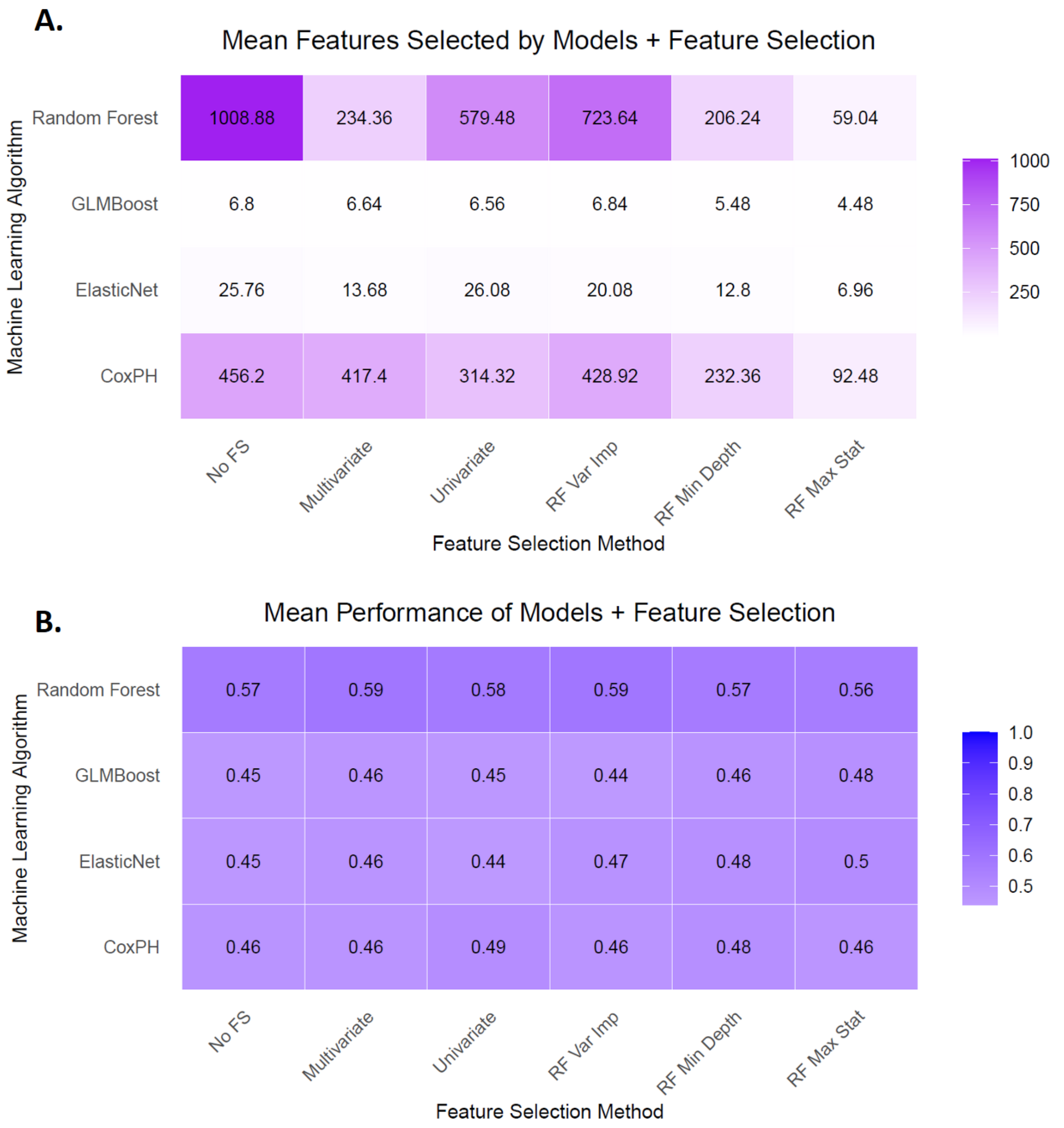

**Supplementary Figure 4:** Distribution of both mir-22 and mir-150 expression across these three age groups.

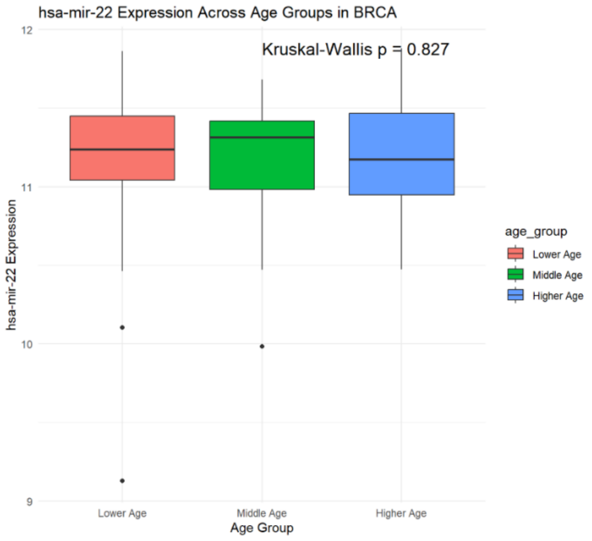


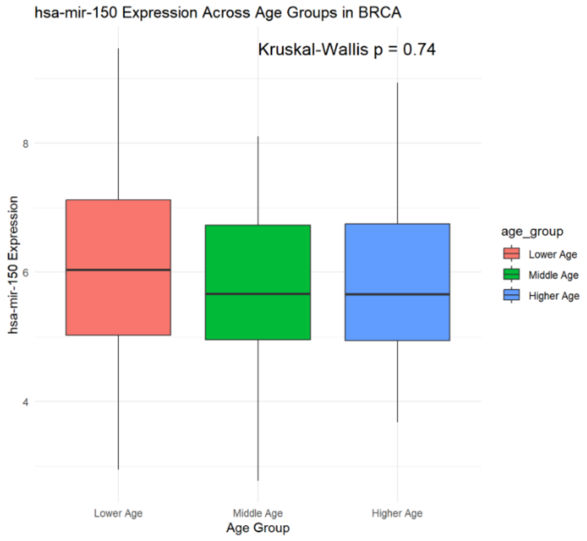


**Supplementary Figure 5:** KM plot showing survival using BRCA miRNA data stratified by age groups.

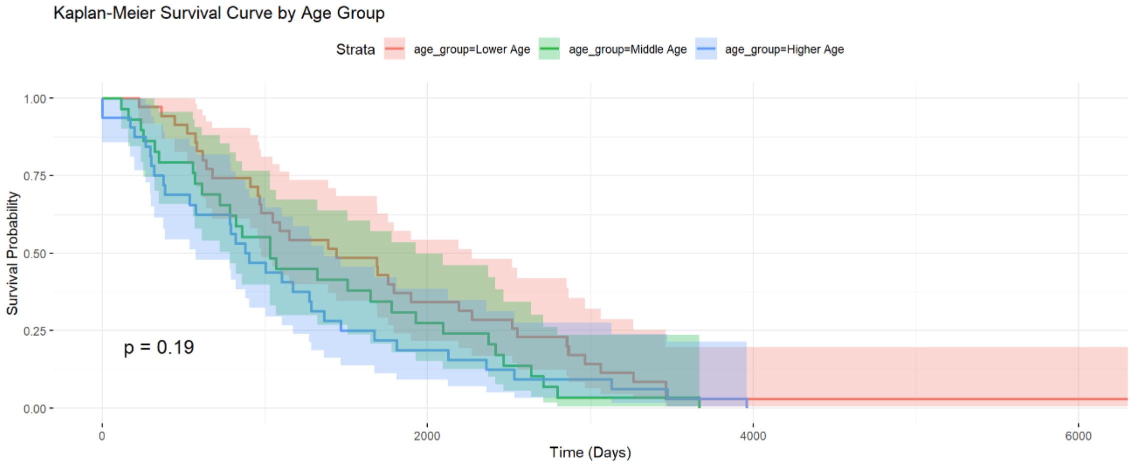
