## Supplementary Tables for "Multi-Omics Prognostic Marker Discovery and Survival Modeling: A Case Study on Pan-Cancer Survival Analyses in Women’s Cancers"

**List of Supplementary Tables**

**Supplementary Table 1.** Comparison of the BRCA dataset, showing the number of features and samples across all modalities before and after the preprocessing steps.

**Supplementary Table 2.** Comparison of preprocessing steps for OV dataset

**Supplementary Table 3**. Comparison of preprocessing steps for CESC dataset

**Supplementary Table 4.** Comparison of preprocessing steps for UCEC dataset

**Supplementary Table 5.** provides a comprehensive summary of the methods applied in our study, including each method's key hyperparameters and the corresponding R package utilized for implementation.

**Supplementary Table 6.** Mean performance with standard deviation and Confidence Intervals from each model and filter methods, using BRCA ME data.

**Supplementary Table 7.** Mean performance with standard deviation and Confidence Intervals from each model and filter methods, using BRCA DM data.

**Supplementary Table 8.** Mean performance with standard deviation and Confidence Intervals from each model and filter methods, using BRCA GE data.

**Supplementary Table 9.** Mean performance with standard deviation and Confidence Intervals from each model and filter methods, using BRCA CNV data.

**Supplementary Table 10.** Overview of the performance of the feature selection pipeline applied to the OV dataset.

**Supplementary Table 11.** Overview of the performance of the feature selection pipeline applied to the CESC dataset.

**Supplementary Table 12.** Overview of the performance of the feature selection pipeline applied to the UCEC dataset.

**Supplementary Table 13**. Comparison between One-Stage and Two Refinement using BRCA dataset

**Supplementary Table 14.** Comparison between One-Stage and Two Refinement using OV dataset

**Supplementary Table 15.** Comparison between One-Stage and Two Refinement using UCEC dataset

**Supplementary Table 16.** Comparison between One-Stage and Two Refinement using CESC dataset

**Supplementary Table 17.** Multi-omics results for BRCA

**Supplementary Table 18.** Multi-omics results for OV

**Supplementary Table 19.** Multi-omics results for CESC

**Supplementary Table 20.** Multi-omics results for UCEC

**Supplementary Table 1.** Comparison of the BRCA dataset, showing the number of features and samples across all modalities before and after the preprocessing steps.

| BRCA | Gene Expression (GE) | Copy Number Variation (CNV) | DNA Methylation (DM) | MiRNA expression (ME) |
|---|---|---|---|---|
| **Features (Original)** | 60660 | 60623 | 485577 | 1881 |
| **Features (Filtered)** | 3033 | 2689 | 1216 | 737 |
| **Samples (Original)** | 1111 | 1050 | 1096 | 1096 |
| **Samples (Filtered)** | 737 | 737 | 737 | 737 |

**Supplementary Table 2.** Comparison of preprocessing steps for OV dataset

| OV | Gene Expression (GE) | Copy Number Variation (CNV) | DNA Methylation (DM) | MiRNA expression (ME) |
|---|---|---|---|---|
| **Features (Original)** | 60660 | 60623 | 485577 | 1881 |
| **Features (Filtered)** | 3033 | 2719 | 1235 | 569 |
| **Samples (Original)** | 421 | 557 | 582 | 490 |
| **Samples (Filtered)** | 395 | 395 | 395 | 395 |

**Supplementary Table 3.** Comparison of preprocessing steps for CESC dataset

| **CESC** | **Gene Expression (GE)** | **Copy Number Variation (CNV)** | **DNA Methylation (DM)** | **MiRNA expression (ME)** |
|---|---|---|---|---|
| **Features (Original)** | 60660 | 60623 | 485577 | 1881 |
| **Features (Filtered)** | 3033 | 2719 | 1217 | 417 |
| **Samples (Original)** | 304 | 294 | 307 | 307 |
| **Samples (Filtered)** | 291 | 291 | 291 | 291 |

**Supplementary Table 4.** Comparison of preprocessing steps for UCEC dataset

| **UCEC** | **Gene Expression (GE)** | **Copy Number Variation (CNV)** | **DNA Methylation (DM)** | **MiRNA expression (ME)** |
|---|---|---|---|---|
| **Features (Original)** | 60660 | 60623 | 485577 | 1881 |
| **Features (Filtered)** | 3033 | 2703 | 1217 | 457 |
| **Samples (Original)** | 553 | 536 | 438 | 545 |
| **Samples (Filtered)** | 420 | 420 | 420 | 420 |

**Supplementary Table 5.** provides a comprehensive summary of the methods applied in our study, including each method's key hyperparameters and the corresponding R package utilized for implementation.

| Method | R package | Function | Hyper-parameters & values |
|---|---|---|---|
| **Learning Algorithms** | | | |
| Cox PH model | *survival* | coxph | |
| Elastic Net | *glmnet* | cv.glmnet | alpha = 0.5, nfolds=5 |
| Gradient boosting with linear models as base learner | *mboost* | glmboost | |
| Random Survival Forests | *ranger* | *ranger* | splitrule = "maxstat",<br>importance = "permutation",<br>mtry: sqrt(#features) -> 100,<br>min.node.size: 50,<br>num.trees=1000 |
| **Feature Selection Methods (filters)** | | | |
| Cox filter | *mlr* | various | |
| RSF variable importance | *randomForestSRC* | rfsrc | ntree = 1000, nsplit = 10<br>mtry = sqrt(#features), nodesize=3 |
| RSF minimal depth | *randomForestSRC* | var.select | method = "md"<br>ntree=1000, nsplit=20, nodesize=5, splitrule="logrank" |
| RSF variable hunting | *randomForestSRC* | var.select | method = "vh", ntree=1000, nodesize=5, splitrule="logrank", nsplit=20, nrep=3, K=10, nstep=1 |

**Supplementary Table 6.** Mean performance with standard deviation and Confidence Intervals from each model and filter methods, using BRCA ME data.

| *C-index* | *No FS* | *Multi-Variate* | *Univariate* | *RF Var Imp* | *RF Min Depth* | *RF Max Stat* |
|---|---|---|---|---|---|---|
| *Random Forest* | 0.63 ± 0.07 (0.60 - 0.65) | 0.62 ± 0.06 (0.59 - 0.64) | 0.63 ± 0.07 (0.61 - 0.66) | 0.63 ± 0.06 (0.61 - 0.65) | 0.62 ± 0.06 (0.59 - 0.64) | 0.56 ± 0.08 (0.53 - 0.59) |
| *GLMBoost* | 0.39 ± 0.06 (0.36 - 0.42) | 0.42 ± 0.07 (0.39 - 0.45) | 0.36 ± 0.07 (0.34 - 0.40) | 0.41 ± 0.07 (0.38 - 0.43) | 0.43 ± 0.08 (0.40 - 0.47) | 0.46 ± 0.08 (0.43 - 0.49) |
| *ElasticNet* | 0.41 ± 0.07 (0.38- 0.44) | 0.43 ± 0.07 (0.41 - 0.46) | 0.39 ± 0.06 (0.37 - 0.41) | 0.41 ± 0.07 (0.38- 0.44) | 0.46 ± 0.07 (0.43 - 0.48) | 0.47 ± 0.08 (0.44 - 0.50) |
| *CoxPH* | 0.48 ± 0.08 (0.45 - 0.52) | 0.47 ± 0.07 (0.44 - 0.49) | 0.40 ± 0.06 (0.37 - 0.42) | 0.50± 0.08 (0.46 - 0.53) | 0.43 ± 0.09 (0.39 - 0.46) | 0.45 ± 0.09 (0.42 - 0.49) |
| *Features Selected* | | | | | | |
| *Random Forest* | 214.4 ± 15.0 (208.6 - 220.3) | 183.7 ± 10.8 (179.5 - 187.9) | 28 ± 8 (24.9 - 31.1) | 162.9 ± 13.3 (157.7 - 168.1) | 64.4 ± 13.5 (59.1 - 69.7) | 9.6 ± 2.3 (8.6 - 10.5) |
| *GLMBoost* | 7.24 ± 1.69 (6.58 - 7.90) | 7 ± 1.6 (6.4 - 7.6) | 7.5 ± 1.8 (6.8 - 8.2) | 7.3 ± 1.8 (6.6 - 8) | 5.9 ± 1.3 (5.4 - 6.4) | 3.8 ± 0.9 (3.4 - 4.1) |
| *ElasticNet* | 18.8 ± 12.2 (14.02- 23.67) | 15.8 ± 16.8 (9.2 - 22.4) | 21.8 ± 3.8 (20.3 - 23.3) | 19.4 ± 12.4 (14.5- 24.2) | 9.9 ± 10.2 (5.9 - 13.9) | 5.1 ± 3.7 (3.6 - 6.5) |
| *CoxPH* | 416.0 ± 0.0 (416.0– 416.0) | 356.4 ± 10.4 (352.3 - 360.5) | 31.9 ± 8.9 (28.4- 35.4) | 300.8 ± 15.8 (294.6 - 307) | 110.8 ± 24.3 (101.2 - 120.3) | 13.9 ± 2.1 (13.1- 14.7) |

**Supplementary Table 7.** Mean performance with standard deviation and Confidence Intervals from each model and filter methods, using BRCA DM data.

| *C-index* | *No FS* | *Multi-Variate* | *Univariate* | *RF Var Imp* | *RF Min Depth* | *RF Max Stat* |
|---|---|---|---|---|---|---|
| *Random Forest* | 0.63 ± 0.06 (0.61 - 0.66) | 0.65 ± 0.06 (0.63 - 0.67) | 0.65 ± 0.06 (0.62 - 0.67) | 0.64 ± 0.06 (0.61 - 0.66) | 0.63 ± 0.06 (0.61 - 0.66) | 0.61 ± 0.06 (0.59 - 0.64) |
| *GLMBoost* | 0.39 ± 0.05 (0.37- 0.42) | 0.41 ± 0.08 (0.38 - 0.45) | 0.36 ± 0.07 (0.33 - 0.39) | 0.41 ± 0.07 (0.38 - 0.44) | 0.39 ± 0.07 (0.37 - 0.42) | 0.44 ± 0.10 (0.40 - 0.48) |
| *ElasticNet* | 0.44 ± 0.06 (0.42 0.47) | 0.48 ± 0.05 (0.46 - 0.50) | 0.38 ± 0.07 (0.35 - 0.41) | 0.44 ± 0.07 (0.42- 0.47) | 0.40 ± 0.08 (0.37 - 0.44) | 0.45 ± 0.08 (0.42 - 0.48) |
| *CoxPH* | 0.51 ± 0.10 (0.47 - 0.55) | 0.46 ± 0.11 (0.42 - 0.51) | 0.45 ± 0.06 (0.43 - 0.48) | 0.49± 0.06 (0.47 - 0.52) | 0.50 ± 0.08 (0.47 - 0.54) | 0.45 ± 0.09 (0.41 - 0.48) |
| *Features Selected* | | | | | | |
| *Random Forest* | 676.56 ± 25.01 (666.76 - 686.36) | 334.88 ± 18.29 (327.71 - 342.05) | 99.28 ± 24.61 (89.63 - 108.93) | 491.16 ± 36.11 (477.00 - 505.32) | 175.76 ± 54.02 (154.59 - 196.93) | 42.24 ± 4.91 (40.32 - 44.16) |
| *GLMBoost* | 9.92 ± 2.27 (9.03 - 10.81) | 9.76 ± 1.85 (9.03 - 10.49) | 9.96 ± 2.26 (9.07 - 10.85) | 9.00 ± 1.71 (8.33 - 9.67) | 7.92 ± 1.93 (7.16 - 8.68) | 6.28 ± 1.34 (5.76 - 6.80) |
| *ElasticNet* | 9.00 ± 8.53 (5.65 - 12.35) | 6.84 ± 12.54 (1.92 - 11.76) | 37.44 ± 12.11 (32.69 - 42.19) | 11.36 ± 10.34 (7.31 - 15.41) | 16.40 ± 7.87 (13.32 - 19.48) | 10.12 ± 9.15 (6.53 - 13.71) |
| *CoxPH* | 600.84 ± 3.13 (599.61 - 602.07) | 593.64 ± 6.43 (591.12 - 596.16) | 123.88 ± 36.65 (109.51 - 138.25) | 598.32 ± 5.76 (596.06 - 600.58) | 284.60 ± 92.28 (248.43 - 320.77) | 59.60 ± 3.96 (58.05 - 61.15) |

**Supplementary Table 8.** Mean performance with standard deviation and Confidence Intervals from each model and filter methods, using BRCA GE data.

| *C-index* | *No FS* | *Multi-Variate* | *Univariate* | *RF Var Imp* | *RF Min Depth* | *RF Max Stat* |
|---|---|---|---|---|---|---|
| *Random Forest* | 0.69 ± 0.06 (0.67 - 0.72) | 0.67 ± 0.06 (0.64 - 0.69) | 0.70 ± 0.06 (0.67 - 0.72) | 0.70 ± 0.05 (0.68 - 0.72) | 0.69 ± 0.06 (0.67 - 0.71) | 0.70 ± 0.06 (0.67 - 0.72) |
| *GLMBoost* | 0.35 ± 0.05 (0.33 - 0.37) | 0.40 ± 0.09 (0.36 - 0.43) | 0.36 ± 0.07 (0.33 - 0.39) | 0.36 ± 0.05 (0.33 - 0.38) | 0.33 ± 0.06 (0.31 - 0.36) | 0.36 ± 0.09 (0.32 - 0.39) |
| *ElasticNet* | 0.34 ± 0.06 (0.32 - 0.37) | 0.43 ± 0.07 (0.40 - 0.46) | 0.33 ± 0.06 (0.31 - 0.36) | 0.37 ± 0.08 (0.34 - 0.40) | 0.34 ± 0.08 (0.30 - 0.37) | 0.36 ± 0.10 (0.32 - 0.39) |
| *CoxPH* | 0.49 ± 0.08 (0.45 - 0.52) | 0.49 ± 0.08 (0.46 - 0.52) | 0.48 ± 0.08 (0.45 - 0.51) | 0.49 ± 0.08 (0.46 - 0.52) | 0.50 ± 0.09 (0.46 - 0.53) | 0.44 ± 0.07 (0.41 - 0.47) |
| *Features Selected* | | | | | | |
| *Random Forest* | 1513.52 ± 41.49 (1497.26 - 1529.78) | 319.76 ± 14.29 (314.16 - 325.36) | 315.08 ± 54.19 (293.84 - 336.32) | 1121.12 ± 42.44 (1104.48 - 1137.76) | 425.52 ± 143.63 (369.22 - 481.82) | 98.92 ± 16.12 (92.60 - 105.24) |
| *GLMBoost* | 10.52 ± 2.18 (9.66 - 11.38) | 8.44 ± 1.39 (7.90 - 8.98) | 10.92 ± 2.20 (10.06 - 11.78) | 11.28 ± 1.86 (10.55 - 12.01) | 8.96 ± 2.26 (8.07 - 9.85) | 7.48 ± 1.08 (7.05 - 7.91) |
| *ElasticNet* | 27.68 ± 19.77 (19.93 - 35.43) | 16.24 ± 14.42 (10.59 - 21.89) | 58.20 ± 22.50 (49.38 - 67.02) | 33.72 ± 21.23 (25.40 - 42.04) | 24.40 ± 20.47 (16.38 - 32.42) | 19.32 ± 12.53 (14.41 - 24.23) |
| *CoxPH* | 602.08 ± 7.42 (599.17 - 604.99) | 591.68 ± 6.62 (589.09 - 594.27) | 432.76 ± 74.45 (403.58 - 461.94) | 606.52 ± 4.05 (604.93 - 608.11) | 521.44 ± 116.64 (475.72 - 567.16) | 158.80 ± 23.04 (149.77 - 167.83) |

**Supplementary Table 9.** Mean performance with standard deviation and Confidence Intervals from each model and filter methods, using BRCA CNV data.

| *C-index* | *No FS* | *Multi-Variate* | *Univariate* | *RF Var Imp* | *RF Min Depth* | *RF Max Stat* |
|---|---|---|---|---|---|---|
| *Random Forest* | 0.57 ± 0.06 (0.55 - 0.59) | 0.59 ± 0.08 (0.56 - 0.62) | 0.58 ± 0.08 (0.55 - 0.61) | 0.59 ± 0.06 (0.57 - 0.61) | 0.57 ± 0.06 (0.55 - 0.59) | 0.56 ± 0.06 (0.54 - 0.59) |
| *GLMBoost* | 0.45 ± 0.07 (0.42 - 0.47) | 0.46 ± 0.10 (0.42 - 0.50) | 0.45 ± 0.07 (0.42 - 0.48) | 0.44 ± 0.11 (0.40 - 0.48) | 0.46 ± 0.09 (0.43 - 0.50) | 0.48 ± 0.08 (0.45 - 0.51) |
| *ElasticNet* | 0.45 ± 0.07 (0.42 - 0.47) | 0.46 ± 0.09 (0.42 - 0.50) | 0.44 ± 0.06 (0.42 - 0.47) | 0.47 ± 0.08 (0.44 - 0.50) | 0.48 ± 0.08 (0.45 - 0.51) | 0.50 ± 0.06 (0.47 - 0.52) |
| *CoxPH* | 0.46 ± 0.08 (0.43 - 0.50) | 0.46 ± 0.10 (0.42 - 0.50) | 0.49 ± 0.07 (0.46 - 0.52) | 0.46 ± 0.09 (0.42 - 0.50) | 0.48 ± 0.09 (0.45 - 0.52) | 0.46 ± 0.09 (0.42 - 0.49) |
| *Features Selected* | | | | | | |
| *Random Forest* | 1008.88 ± 87.06 (974.75 - 1043.01) | 34.36 ± 21.40 (225.97 - 242.75) | 579.48 ± 150.25 (520.58 - 638.38) | 723.64 ± 81.89 (691.54 - 755.74) | 206.24 ± 96.75 (168.31 - 244.17) | 59.04 ± 14.25 (53.45 - 64.63) |
| *GLMBoost* | 6.80 ± 1.58 (6.18 - 7.42) | 6.64 ± 1.58 (6.02 - 7.26) | 6.56 ± 1.39 (6.02 - 7.10) | 6.84 ± 1.77 (6.15 - 7.53) | 5.48 ± 1.45 (4.91 - 6.05) | 4.48 ± 1.50 (3.89 - 5.07) |
| *ElasticNet* | 25.76 ± 11.34 (21.31 - 30.21) | 13.68 ± 5.90 (11.37 - 15.99) | 26.08 ± 8.33 (22.82 - 29.34) | 20.08 ± 12.88 (15.03 - 25.13) | 12.80 ± 6.78 (10.14 - 15.46) | 6.96 ± 4.53 (5.18 - 8.74) |
| *CoxPH* | 456.2 ± 8.26 (452.96 - 459.44) | 417.4 ± 9.95 (413.50 - 421.30) | 314.32 ± 43.98 (297.08 - 331.56) | 428.92 ± 13.17 (423.76 - 434.08) | 232.36 ± 101.33 (192.64 - 272.08) | 92.48 ± 11.68 (87.90 - 97.06) |

**Supplementary Table 10.** Overview of the performance of the feature selection pipeline applied to the OV dataset.

| OV | Omics | None | CV |
|---|---|---|---|
| **CoxPH (1)** | GE | 0.4963 ± 0.0402 (0.4230 - 0.5402) | 0.4045 ± 0.0384 (0.3538 - 0.4437) |
| | ME | 0.4679 ± 0.0355 (0.4114 - 0.5161) | 0.4821 ± 0.0451 (0.4217 - 0.5535) |
| | DM | 0.4589 ± 0.0309 (0.4059 - 0.5031) | 0.4569 ± 0.0325 (0.4141 - 0.5088) |
| | CNV | 0.4553 ± 0.0415 (0.4007 - 0.5116) | 0.5400 ± 0.0478 (0.4543 - 0.6159) |
| **Ranger (2)** | GE | **0.4835 ± 0.0358 (0.4388 - 0.5366)** | **0.5331 ± 0.0263 (0.4993 - 0.5690)** |
| | ME | **0.5789 ± 0.0594 (0.4867 - 0.6830)** | **0.6037 ± 0.0492 (0.5283 - 0.6746)** |
| | DM | **0.5853 ± 0.0362 (0.5072 - 0.6119)** | **0.5640 ± 0.0509 (0.4880 - 0.6248)** |
| | CNV | **0.4620 ± 0.0490 (0.4092 - 0.5229)** | **0.4487 ± 0.0494 (0.3633 - 0.5012)** |
| **ElasticNet (3)** | GE | 0.4735 ± 0.0397 (0.4135 - 0.5270) | 0.4404 ± 0.0335 (0.3914 - 0.5015) |
| | ME | 0.4625 ± 0.0472 (0.4025 - 0.5398) | 0.4422 ± 0.0235 (0.4102 - 0.4855) |
| | DM | 0.4778 ± 0.0326 (0.4412 - 0.5389) | 0.4515 ± 0.0250 (0.4122 - 0.4879) |
| | CNV | 0.5037 ± 0.0320 (0.4702 - 0.5647) | 0.4939 ± 0.0299 (0.4562 - 0.5408) |
| **GLMBoost (4)** | GE | 0.4426 ± 0.0326 (0.4053 - 0.4986) | 0.5000 ± 0.0000 (0.5 - 0.5) |
| | ME | 0.4472 ± 0.0419 (0.3828 - 0.5168) | 0.4240 ± 0.0326 (0.3793 - 0.4768 |
| | DM | 0.5374 ± 0.0382 (0.4721 - 0.5892) | 0.4525 ± 0.0530 (0.3680 - 0.5115) |
| | CNV | 0.5000 ± 0.0000 (0.5000 - 0.5000) | 0.5000 ± 0.0000 (0.5000 - 0.5000) |

**Supplementary Table 11.** Overview of the performance of the feature selection pipeline applied to the CESC dataset.

| CESC | Omics | None | CV |
|---|---|---|---|
| **CoxPH (1)** | GE | 0.3785 ± 0.0864 (0.2507 - 0.4819) | 0.3497 ± 0.1181 (0.2111 - 0.5838) |
| | ME | 0.6596 ± 0.0775 (0.5395 - 0.7712) | 0.3191 ± 0.0440 (0.2588 - 0.3742) |
| | DM | 0.4962 ± 0.0600 (0.4142 - 0.5715) | 0.6063 ± 0.0641 (0.4941 - 0.7098) |
| | CNV | 0.5646 ± 0.0813 (0.4550 - 0.6842) | 0.4809 ± 0.0903 (0.3383 - 0.5981) |
| **Ranger (2)** | GE | **0.7195 ± 0.0531 (0.6226 - 0.7848)** | **0.6024 ± 0.0533 (0.5463 - 0.6963)** |
| | ME | **0.5911 ± 0.0820 (0.4328 - 0.6862)** | **0.5640 ± 0.0827 (0.4064 - 0.6401)** |
| | DM | **0.5917 ± 0.0647 (0.4839 - 0.6802)** | **0.5554 ± 0.0736 (0.4484 - 0.6683)** |
| | CNV | **0.4410 ± 0.0439 (0.3788 - 0.5066)** | **0.4432 ± 0.0714 (0.3527 - 0.5675)** |
| **ElasticNet (3)** | GE | 0.2533 ± 0.0390 (0.2002 - 0.3017) | 0.3573 ± 0.0840 (0.2172 - 0.4491) |
| | ME | 0.3452 ± 0.0719 (0.2204 - 0.4400) | 0.2513 ± 0.0383 (0.2071 - 0.3209) |
| | DM | 0.4512 ± 0.0596 (0.3665 - 0.5538) | 0.4991 ± 0.1029 (0.3728 - 0.6604) |
| | CNV | 0.6007 ± 0.0959 (0.4304 - 0.7526) | 0.4485 ± 0.0373 (0.3959 - 0.5174) |
| **GLMBoost (4)** | GE | 0.2967 ± 0.0652 (0.1834 - 0.3838) | 0.3656 ± 0.1444 (0.1865 - 0.5999) |
| | ME | 0.3264 ± 0.0843 (0.1886 - 0.4590) | 0.2778 ± 0.0775 (0.1988 - 0.4351) |
| | DM | 0.4922 ± 0.0597 (0.3923 - 0.5888) | 0.5118 ± 0.0601 (0.4222 - 0.5989) |
| | CNV | 0.5534 ± 0.0719 (0.4389 - 0.6450) | 0.4312 ± 0.0351 (0.3902 - 0.4880) |

**Supplementary Table 12.** Overview of the performance of the feature selection pipeline applied to the UCEC dataset.

| UCEC | Omics | None | CV |
|---|---|---|---|
| **CoxPH (1)** | GE | 0.4924 ± 0.0760 (0.3782 - 0.6136) | 0.5880 ± 0.1066 (0.4167 - 0.7327) |
| | ME | 0.4324 ± 0.0657 (0.3363 - 0.5207) | 0.3720 ± 0.0607 (0.3002 - 0.4637) |
| | DM | 0.4171 ± 0.0724 (0.2977 - 0.5022) | 0.5049 ± 0.0523 (0.4208 - 0.5874) |
| | CNV | 0.3980 ± 0.1141 (0.2506 - 0.5538) | 0.4715 ± 0.1221 (0.2800 - 0.6330) |
| **Ranger (2)** | GE | **0.5710 ± 0.0694 (0.4599 - 0.6662)** | **0.5526 ± 0.0488 (0.4650 - 0.6121)** |
| | ME | **0.6929 ± 0.0518 (0.6004 - 0.7578)** | **0.7752 ± 0.0399 (0.7123 - 0.8284)** |
| | DM | **0.7628 ± 0.0401 (0.7095 - 0.8244)** | **0.7352 ± 0.0428 (0.6918 - 0.8083)** |
| | CNV | **0.7364 ± 0.0490 (0.6677 - 0.8066)** | **0.7678 ± 0.0535 (0.6535 - 0.8157)** |
| **ElasticNet (3)** | GE | 0.4332 ± 0.0711 (0.3325 - 0.5435) | 0.4242 ± 0.0719 (0.3230 - 0.5365) |
| | ME | 0.4538 ± 0.0787 (0.3586 - 0.5921) | 0.3291 ± 0.0703 (0.2182 - 0.4159) |
| | DM | 0.3056 ± 0.0638 (0.2067 - 0.3894) | 0.2436 ± 0.0441 (0.1745 - 0.3128) |
| | CNV | 0.3653 ± 0.0707 (0.2733 - 0.4646) | 0.3813 ± 0.0630 (0.3078 - 0.4912) |
| **GLMBoost (4)** | GE | 0.4394 ± 0.0610 (0.3767 - 0.5398) | 0.4130 ± 0.0659 (0.3113 - 0.5003) |
| | ME | 0.5000 ± 0.0000 (0.5000 - 0.5000) | 0.4123 ± 0.0671 (0.2946 - 0.4937) |
| | DM | 0.2998 ± 0.0654 (0.1819 - 0.3652) | 0.2422 ± 0.0442 (0.1687 - 0.3094) |
| | CNV | 0.3678 ± 0.0424 (0.3027 - 0.4394) | 0.3471 ± 0.0713 (0.2332 - 0.4341) |

**Supplementary Table 13.** Comparison between One-Stage and Two Refinement using BRCA dataset

| **Modality combinations** | **ME + GE** | **ME + DM** | **GE + DM** | **GE + CNV** | **ME + CNV** | **DM + CNV** | **DM + GE + ME** | **DM + GE + CNV** | **DM + ME + CNV** | **ME + GE + CNV** | **ME + GE + CNV+ DM** |
|---|---|---|---|---|---|---|---|---|---|---|---|
| C-index of One-Stage Refinement | 0.69 ± 0.004 (0.69 - 0.70) | 0.70 ± 0.005 (0.70 - 0.71) | 0.70 ± 0.04 (0.67 - 0.73) | 0.70 ± 0.04 (0.68 - 0.74) | 0.68 ± 0.03 (0.65 - 0.70) | 0.63 ± 0.04 (0.60 - 0.66) | 0.69 ± 0.05 (0.66 - 0.73) | 0.70 ± 0.05 (0.67 - 0.74) | 0.69 ± 0.05 (0.66 - 0.73) | 0.70 ± 0.05 (0.67 - 0.74) | **0.71 ± 0.05 (0.68 - 0.75)** |
| C-index of Two-Stage Refinement | 0.74 ± 0.03 (0.71 - 0.77) | 0.69 ± 0.03 (0.66 - 0.71) | 0.72 ± 0.04 (0.70 - 0.75) | 0.71 ± 0.04 (0.67 - 0.74) | 0.72 ± 0.04 (0.69 - 0.75) | 0.68 ± 0.06 (0.63 - 0.73) | 0.73 ± 0.03 (0.71 - 0.75) | 0.70 ± 0.03 (0.68 - 0.73) | **0.77 ± 0.03 (0.73 - 0.80)** | 0.75 ± 0.03 (0.71 - 0.78) | 0.71 ± 0.05 (0.68 - 0.75) |

**Supplementary Table 14.** Comparison between One-Stage and Two Refinement using OV dataset

| **Modality combinations** | **ME + GE** | **ME + DM** | **GE + DM** | **GE + CNV** | **ME + CNV** | **DM + CNV** | **DM + GE + ME** | **DM + GE + CNV** | **DM + ME + CNV** | **ME + GE + CNV** | **ME + GE + CNV + DM** |
|---|---|---|---|---|---|---|---|---|---|---|---|
| C-index of One-Stage Refinement | 0.59 ± 0.03 (0.57 - 0.61) | **0.62 ± 0.03 (0.60 - 0.64)** | 0.59 ± 0.03 (0.57 - 0.61) | 0.59 ± 0.04 (0.56 - 0.62) | 0.60 ± 0.02 (0.59 - 0.63) | 0.59 ± 0.03 (0.57 - 0.61) | 0.61 ± 0.03 (0.59 - 0.64) | 0.58 ± 0.02 (0.57 - 0.60) | 0.61 ± 0.01 (0.60 - 0.63) | 0.61 ± 0.03 (0.59 - 0.63) | 0.61 ± 0.03 (0.59 - 0.62) |
| C-index of Two-Stage Refinement | 0.63 ± 0.02 (0.62 - 0.65) | 0.64 ± 0.02 (0.62 - 0.66) | 0.61 ± 0.02 (0.60 - 0.63) | 0.59 ± 0.06 (0.55 - 0.63) | 0.61 ± 0.03 (0.59 - 0.63) | 0.59 ± 0.02 (0.57 - 0.61) | **0.65 ± 0.02 (0.63 - 0.67)** | 0.60 ± 0.03 (0.58 - 0.63) | 0.60 ± 0.03 (0.58 - 0.62) | 0.63 ± 0.03 (0.60 - 0.65) | 0.63 ± 0.03 (0.61 - 0.65) |

**Supplementary Table 15.** Comparison between One-Stage and Two Refinement using UCEC dataset

| **Modality combinations** | **ME + GE** | **ME + DM** | **GE + DM** | **GE + CNV** | **ME + CNV** | **DM + CNV** | **DM + GE + ME** | **DM + GE + CNV** | **DM + ME + CNV** | **ME + GE + CNV** | **ME + GE + CNV + DM** |
|---|---|---|---|---|---|---|---|---|---|---|---|
| C-index of One-Stage Refinement | 0.69 ± 0.05 (0.67 - 0.73) | 0.69 ± 0.06 (0.65 - 0.74) | **0.73 ± 0.06 (0.69 - 0.77)** | 0.59 ± 0.04 (0.56 - 0.62) | 0.67 ± 0.05 (0.64 - 0.70) | 0.70 ± 0.04 (0.66 - 0.73) | 0.70 ± 0.03 (0.68 - 0.72) | 0.71 ± 0.03 (0.69 - 0.74) | 0.69 ± 0.04 (0.66 - 0.72) | 0.70 ± 0.03 (0.68 - 0.72) | 0.69 ± 0.07 (0.64 - 0.74) |
| C-index of Two-Stage Refinement | 0.70 ± 0.05 (0.67 - 0.74) | 0.73 ± 0.05 (0.69 - 0.76) | 0.71 ± 0.05 (0.68 - 0.75) | 0.68 ± 0.06 (0.64 - 0.72) | 0.70 ± 0.02 (0.68 - 0.72) | 0.70 ± 0.04 (0.67 - 0.73) | 0.72 ± 0.05 (0.69 - 0.76) | 0.72 ± 0.05 (0.69 - 0.76) | **0.76 ± 0.03 (0.72 - 0.80)** | 0.70 ± 0.06 (0.66 - 0.75) | 0.71 ± 0.03 (0.68 - 0.74) |

**Supplementary Table 16.** Comparison between One-Stage and Two Refinement using CESC dataset

| **Modality combinations** | **ME + GE** | **ME + DM** | **GE + DM** | **GE + CNV** | **ME + CNV** | **DM + CNV** | **DM + GE + ME** | **DM + GE + CNV** | **DM + ME + CNV** | **ME + GE + CNV** | **ME + GE + CNV+ DM** |
|---|---|---|---|---|---|---|---|---|---|---|---|
| C-index of One-Stage Refinement | 0.69 ± 0.04 (0.67 - 0.72) | **0.70 ± 0.05 (0.67 - 0.74)** | 0.62 ± 0.06 (0.57 - 0.66) | 0.66 ± 0.04 (0.64 - 0.69) | 0.69 ± 0.03 (0.67 - 0.71) | 0.62 ± 0.03 (0.59 - 0.65) | 0.68 ± 0.04 (0.65 - 0.72) | 0.63 ± 0.05 (0.60 - 0.68) | 0.69 ± 0.06 (0.65 - 0.74) | 0.68 ± 0.03 (0.66 - 0.70) | 0.69 ± 0.06 (0.66 - 0.74) |
| C-index of Two-Stage Refinement | 0.72 ± 0.05 (0.69 - 0.76) | 0.76 ± 0.05 (0.72 - 0.79) | 0.71 ± 0.06 (0.67 - 0.75) | 0.69 ± 0.06 (0.64 - 0.73) | 0.71 ± 0.07 (0.66 - 0.76) | 0.65 ± 0.04 (0.63 - 0.68) | 0.74 ± 0.06 (0.70 - 0.78) | 0.73 ± 0.04 (0.70 - 0.76) | **0.80 ± 0.03 (0.76 - 0.83)** | 0.71 ± 0.04 (0.68 - 0.73) | 0.75 ± 0.03 (0.73 - 0.77) |

**Supplementary Table 17.** *Multi-omics results for BRCA*

| Modality combinations | ME + GE | ME + DM | GE + DM | GE + CNV | ME + CNV | DM + CNV | DM + GE + ME | DM + GE + CNV | DM + ME + CNV | ME + GE + CNV | ME + GE + CNV + DM |
|---|---|---|---|---|---|---|---|---|---|---|---|
| C-index of Two-Stage Refinement | 0.74 | 0.69 | 0.72 | 0.71 | 0.72 | 0.68 | 0.73 | 0.70 | **0.77** | 0.75 | 0.71 |

| Omics | Features |
|---|---|
| **DM** | cg15520279, cg18087514, cg02776251, cg24262376, cg01671575, cg14717170, cg20786074, cg09656934, cg20308679, cg10634551, cg12626411, cg01664666, cg07837085, cg20261167, cg15147516, cg08205865, cg25583174, cg12958813, cg15761405, cg25167447, cg04574507, cg20676475, cg11902458, cg00807586, cg01637734, cg02836529, cg26583078, cg16232126, cg17214107, cg08422599, cg09952204, cg10978355, cg05619712, cg21137417, cg09563216, cg02983451, cg15077070, cg11657808, cg20311501, cg05275752, cg02431964, cg17453778, cg18335243, cg18182399, cg19616230, cg10777851, cg03679305, cg23300372, cg01612158, cg12815916, cg03614513, cg21591742, cg18006568, cg27403635, cg23282674, cg23557926, cg22176895, cg13320626, cg06933072, cg18110483, cg22359606, cg09220361, cg13847070, cg22855405, cg07038400, cg13986130, cg07354440, cg01114088, cg18794577, cg16557944, cg18482268, cg11428724, cg20613889, cg00995327, cg02774439, cg01216369, cg24512973, cg17701886, cg16516400, cg11819637, cg09478478, cg05200628, cg12105450, cg17525406, cg04098585, cg08474603, cg00884221, cg25462303, cg17105014, cg17542385, cg17020834, cg07548313, cg01868128, cg14925024, cg05768141, cg17108819, cg08826839, cg05909475, cg25057743, cg25631352, cg08047907, cg10861599, cg08658594, cg10691387, cg18277754, cg08578023, cg12880658, cg17281600, cg08690031, cg19228118, cg10031651, cg03702236, cg08057475, cg22605643, cg25437385, cg03852144, cg21948655, cg03329572, cg06168050, cg06274159, cg23363832, cg10370591, cg11724759, cg05248781, cg10774440, cg07664856, cg21972382, cg20535085, cg07694025, cg24928687, cg12864235, cg20579480, cg14419187, cg06043114, cg19884658, cg17430393, cg19526600, cg00498604, cg03199651, cg11976166, cg16139316, cg06940792, cg00567749, cg10946435, cg21991396, cg18908499, cg01367992, cg26117023 |
| **ME** | hsa.mir.31, hsa.mir.30a, hsa.mir.22, hsa.mir.150, hsa.mir.4742, hsa.mir.221 |
| **CNV** | ADAM2, AC139365.1, RAB11FIP1, CYP4F44P, AC239800.2, U3, RNU1.124P, HSPA8P13, POMK, AC110275.1, AC139365.2, AC123767.1, RNU6.356P, AC103726.1, AC067817.2, ZNF703, SLC20A2, RPS20P22, AC048387.1, MIR1204, ADAM3A, RN7SL709P, AC118650.1, SNORD65B, SFRP1, MIR548AO, AC091182.1 |

**Supplementary Table 18.** *Multi-omics results for OV*

| **Modality combinations** | **ME + GE** | **ME + DM** | **GE + DM** | **GE + CNV** | **ME + CNV** | **DM + CNV** | **DM + GE + ME** | **DM + GE + CNV** | **DM + ME + CNV** | **ME + GE + CNV** | **ME + GE + CNV + DM** |
|---|---|---|---|---|---|---|---|---|---|---|---|
| C-index of Two-Stage Refinement | 0.63 | 0.64 | 0.61 | 0.59 | 0.61 | 0.59 | **0.65** | 0.60 | 0.60 | 0.63 | 0.63 |

| **Omics** | **Features** |
|---|---|
| **DM** | cg04533291, cg03874199, cg27342801, cg20676475, cg06101324 |
| **ME** | hsa.mir.1301, hsa.mir.3200, hsa.let.7a.1, hsa.let.7a.3, hsa.mir.135b, hsa.mir.139 |

**Supplementary Table 19.** *Multi-omics results for CESC*

| **Modality combinations** | **ME + GE** | **ME + DM** | **GE + DM** | **GE + CNV** | **ME + CNV** | **DM + CNV** | **DM + GE + ME** | **DM + GE + CNV** | **DM + ME + CNV** | **ME + GE + CNV** | **ME + GE + CNV + DM** |
|---|---|---|---|---|---|---|---|---|---|---|---|
| C-index of Two-Stage Refinement | 0.72 | 0.76 | 0.71 | 0.69 | 0.71 | 0.65 | 0.74 | 0.73 | **0.80** | 0.71 | 0.75 |

| **Omics** | **Features** |
|---|---|
| **DM** | cg25514503, cg16607065, cg01612158, cg12958813 |
| **ME** | hsa.mir.502, hsa.mir.101.2, hsa.mir.140, hsa.mir.150, hsa.mir.500b, hsa.mir.1306, hsa.mir.142, hsa.mir.204, hsa.mir.144, hsa.mir.188, hsa.mir.155, hsa.mir.205, hsa.mir.335, hsa.mir.148a, hsa.mir.196a.1, hsa.mir.151a, hsa.mir.193a, hsa.let.7e |
| **CNV** | FAM91A2P, RNU6.488P |

**Supplementary Table 20.** *Multi-omics results for UCEC*

| Modality combinations | ME + GE | ME + DM | GE + DM | GE + CNV | ME + CNV | DM + CNV | DM + GE + ME | DM + GE + CNV | DM + ME + CNV | ME + GE + CNV | ME + GE + CNV + DM |
|---|---|---|---|---|---|---|---|---|---|---|---|
| C-index of Two-Stage Refinement | 0.70 | 0.73 | 0.71 | 0.68 | 0.70 | 0.70 | 0.72 | 0.72 | **0.76** | 0.70 | 0.71 |

| Omics | Features |
|---|---|
| **DM** | cg11009736, cg12177677, cg17525406, cg22751696, cg06275635, cg13968061, cg19884600, cg10849854, cg16607065, cg06096336, cg14419187, cg22605643, cg04391111, cg12417466, cg26353877, cg05368341, cg14458615, cg04660410, cg23477967, cg15175266, cg08763351, cg08804892, cg13901526, cg14991487, cg09478478, cg06490988, cg10885338, cg11657808, cg21376883, cg02192965, cg25596297, cg18277754, cg25115460, cg10523019, cg25856383, cg06856528, cg12105450, cg01426743, cg14696870, cg22338307, cg07548313 |
| **ME** | hsa.mir.149, hsa.mir.222, hsa.mir.598, hsa.mir.335, hsa.mir.150, hsa.mir.1224, hsa.let.7a.3, hsa.mir.2355, hsa.mir.31, hsa.mir.146a, hsa.let.7a.1, hsa.mir.193b, hsa.mir.142, hsa.mir.140, hsa.mir.187, hsa.mir.155, hsa.mir.3131, hsa.mir.3614, hsa.mir.1976, hsa.mir.1468, hsa.mir.152, hsa.mir.1249, hsa.mir.143, hsa.let.7a.2, hsa.mir.181b.1, hsa.mir.374a, hsa.mir.1266, hsa.mir.153.2, hsa.let.7f.2, hsa.mir.127, hsa.mir.144, hsa.mir.29a, hsa.mir.194.2, hsa.mir.30e, hsa.mir.29b.2, hsa.let.7f.1, hsa.mir.10b, hsa.mir.181b.2, hsa.mir.22, hsa.mir.16.1, hsa.mir.203b, hsa.mir.191, hsa.mir.1287, hsa.let.7g, hsa.mir.192, hsa.mir.1226, hsa.mir.28, hsa.mir.30c.1, hsa.mir.25, hsa.mir.10a, hsa.mir.301b, hsa.mir.135b, hsa.mir.27b, hsa.mir.133a.1, hsa.mir.133b, hsa.let.7d, hsa.mir.1343, hsa.mir.214, hsa.mir.199a.2, hsa.mir.125b.1, hsa.mir.29b.1, hsa.mir.194.1, hsa.mir.181d, hsa.mir.129.1, hsa.mir.130a, hsa.mir.30a, hsa.mir.20b, hsa.mir.199a.1, hsa.mir.133a.2, hsa.mir.196a.1, hsa.mir.129.2, hsa.mir.223, hsa.mir.148a, hsa.mir.125b.2 |
| **CNV** | MIR1204, Y_RNA, AL499627.1, AC104653.2, AC017074.1, TERC, RSL24D1P6, AC022634.1, MIR8083, U3, AC104640.1, RNVU1.4, AC022296.2, Metazoa_SRP |
